# Prognostic Insights from Longitudinal Multicompartment Study of Host-Microbiota Interactions in Critically Ill Patients

**DOI:** 10.1101/2023.09.25.23296086

**Authors:** Georgios D. Kitsios, Khaled Sayed, Adam Fitch, Haopu Yang, Noel Britton, Faraaz Shah, William Bain, John W. Evankovich, Shulin Qin, Xiaohong Wang, Kelvin Li, Asha Patel, Yingze Zhang, Josiah Radder, Charles Dela Cruz, Daniel A Okin, Ching-Ying Huang, Daria van Tyne, Panayiotis V. Benos, Barbara Methé, Peggy Lai, Alison Morris, Bryan J. McVerry

## Abstract

Critical illness can disrupt the composition and function of the microbiome, yet comprehensive longitudinal studies are lacking. We conducted a longitudinal analysis of oral, lung, and gut microbiota in a large cohort of 479 mechanically ventilated patients with acute respiratory failure. Progressive dysbiosis emerged in all three body compartments, characterized by reduced alpha diversity, depletion of obligate anaerobe bacteria, and pathogen enrichment. Clinical variables, including chronic obstructive pulmonary disease, immunosuppression, and antibiotic exposure, shaped dysbiosis. Notably, of the three body compartments, unsupervised clusters of lung microbiota diversity and composition independently predicted survival, transcending clinical predictors, organ dysfunction severity, and host-response sub-phenotypes. These independent associations of lung microbiota may serve as valuable biomarkers for prognostication and treatment decisions in critically ill patients. Insights into the dynamics of the microbiome during critical illness highlight the potential for microbiota-targeted interventions in precision medicine.

## Introduction

Microbiota play a critical role in maintaining homeostasis and overall health. However, during critical illness, such as acute respiratory failure (ARF), microbial communities can be severely disrupted.^1,2^ Such disruptions, characterized by deviations from a healthy microbial composition and diversity, may occur early in the hospital stay and have been associated with worse clinical outcomes.^3–5^ Previous research has primarily focused on cross-sectional analyses of microbiota within individual body sites, neglecting potential interactions between different compartments and the longitudinal evolution of microbial communities. Moreover, the influence of patient-level factors and therapeutic interventions, including antimicrobial therapies, on the microbiome of critically ill patients remains poorly understood, partly due to limitations of scale in studies published to date.

Precision medicine approaches in ARF have predominantly focused on host factors.^6^ For instance, identifying distinct subphenotypes based on patterns of host response biomarkers measured in plasma samples (hyper-vs. hypo-inflammatory) has demonstrated prognostic value.^7–9^ Hyperinflammatory patients exhibit elevated levels of injury and inflammation biomarkers, more severe organ dysfunction, worse prognosis, and may have distinct responses to treatments.^8^ However, the role of respiratory or intestinal microbiota in modulating host responses and their contributions to defined subphenotypes are still not well understood.

Furthermore, limited data are available regarding the potential influence of respiratory microbiota on systemic host responses measured in plasma or localized inflammation within the lungs.^10^ To advance precision medicine approaches that take into account the microbial side of the critically ill host, it is crucial to understand the dynamics of the microbiome and its relationship with host biological factors, clinical diagnoses, and therapeutic interventions in critical illness.

To address these knowledge gaps, we conducted a longitudinal assessment of the microbiome in a large cohort of 479 ARF patients, specifically focusing on three key body sites: the oral cavity, lungs, and gut. By integrating bacterial and fungal community profiles with host response biomarkers measured in plasma and lower respiratory tract (LRT) samples, we examined the temporal associations between patient-level factors and therapeutic interventions on microbial communities. We derived unsupervised clusters of microbiota and determined their associations with host-response subphenotypes and clinical outcomes. Finally, we validated our findings in two separate cohorts with a total of 146 patients with COVID-19-associated ARF.

## Results

### Cohort Description

We performed discovery analyses in a cohort of 479 patients with ARF who received invasive mechanical ventilation (IMV) via endotracheal intubation in UPMC Intensive Care Units (ICUs) (**UPMC-ARF cohort**), and then independent validation analyses in two cohorts of critically ill patients with COVID-19 pneumonia (49 patients at UPMC **[UPMC-COVID cohort],** and 97 patients at Massachusetts General Hospital ICUs, **MGH-COVID** cohort).

In the UPMC-ARF cohort, we enrolled patients with non-COVID etiologies of ARF between March 2015 and June 2022. We collected baseline research biospecimens within 72hrs from intubation, including blood for separation of plasma, oropharyngeal swabs (oral samples), endotracheal aspirates (ETA) collected for research or excess bronchoalveolar lavage fluid (BALF) from clinical bronchoscopy (lung samples), and rectal swabs or stool (gut samples).^3,11,12^ We repeated research biospecimen sampling between days 3-6 (middle interval) and days 7-12 (late interval) post-enrollment for subjects who remained in the ICU. We extracted DNA and performed next-generation sequencing (bacterial 16S rRNA gene sequencing [16S-Seq] for all available samples; fungal Internal Transcribed Spacer sequencing [ITS-Seq] targeting the regions 1 and 2 of the ITS rRNA gene, and Nanopore DNA metagenomics for a subset of samples) to profile microbiota in the oral, lung and gut communities, respectively.^3,12,13^ We measured biomarker proteins in plasma samples and ETA/BALF supernatants with Luminex panels to profile systemic and regional (lung) host responses.^7,10^

Patients had a median (interquartile range) age of 59.6 (46.7-68.7) years, 54.4% were men and 90.2% were whites (Table 1). At the time of enrollment, 25.0% of patients were diagnosed with Acute Respiratory Distress Syndrome (ARDS per the Berlin definition^14^) and 39.8% with pneumonia, 86.8% were receiving systemic antibiotics, and 64.8% received corticosteroids for various indications. By 60 days, 26.9% of patients had died. Among the 350 patients who survived hospitalization, 48.8% were discharged to their home, with the remainder requiring additional longer-term care.

**Table 1:** Baseline characteristics of enrolled mechanically ventilated patients in the UPMC-ARF cohort, stratified by 60-day mortality. We compared continuous variables with non-parametric Wilcoxon tests and categorical variables with Fisher’s exact tests between the three groups. Statistically significant differences (p<0.05) are highlighted in bold.

|  | All | Survivors | Non-Survivors | p |
| --- | --- | --- | --- | --- |
| N | 479 | 350 | 129 |  |
| Age, years (median [IQR]) | 59.6 [46.7, 68.7] | 57.1 [44.1, 67.1] | 65.3 [55.8, 72.2] | <0.01 |
| Men, n (%) | 256 (54.4) | 180 (52.6) | 76 (58.9) | 0.26 |
| Whites, n (%) | 425 (90.2) | 307 (89.8) | 118 (91.5) | 0.67 |
| BMI (median [IQR]) | 29.4 [25.5, 36.0] | 29.6 [25.5, 35.7] | 28.6 [25.3, 36.6] | 0.98 |
| COPD, n (%) | 104 (22.1) | 75 (21.9) | 29 (22.5) | 1.00 |
| Diabetes, n (%) | 168 (35.7) | 122 (35.7) | 46 (35.7) | 1.00 |
| Alcohol use, n (%) | 84 (17.9) | 60 (17.5) | 24 (18.9) | 0.84 |
| Immunosuppression, n (%) | 105 (22.3) | 71 (20.8) | 34 (26.4) | 0.24 |
| ARDS, n (%) | 117 (25.2) | 81 (24.0) | 36 (28.1) | 0.23 |
| WBC (median [IQR]) | 12.0 [8.7, 16.8] | 11.4 [8.1, 15.8] | 14.4 [10.1, 18.7] | <0.01 |
| Creatinine (median [IQR]) | 1.2 [0.8, 2.3] | 1.1 [0.8, 2.0] | 1.6 [0.9, 2.5] | 0.01 |
| Plateau Pressure (median [IQR]) | 20.0 [16.0, 25.0] | 19.0 [16.0, 24.0] | 22.0 [18.0, 27.0] | <0.01 |
| PaO<sub>2</sub>:FiO<sub>2</sub> ratio (median [IQR]) | 164.0 [117.0, 206.0] | 168.0 [121.5, 211.0] | 157.0 [108.0, 205.0] | 0.04 |
| SOFA scores (median [IQR]) | 6.0 [4.0, 9.0] | 6.0 [4.0, 8.0] | 8.0 [5.0, 10.0] | <0.01 |
| LIPS score (median [IQR]) | 5.5 [4.0, 6.5] | 5.0 [4.0, 6.5] | 6.0 [5.0, 7.5] | <0.01 |
| Hypoinflammatory subphenotype, n (%) | 344 (75.6) | 254 (77.4) | 90 (70.9) | 0.18 |
| VFD (median [IQR]) | 22.0 [13.0, 25.0] | 23.0 [20.0, 25.2] | 0.0 [0.0, 19.0] | <0.01 |

Abbreviations: IQR: Interquartile Range; BMI: body mass index; COPD: chronic obstructive pulmonary disease, LIPS: lung injury prediction score; WBC: white blood cell count; PaO<sub>2</sub>: partial pressure of arterial oxygen; FiO<sub>2</sub>: Fractional inhaled concentration of oxygen; SOFA: sequential organ failure assessment; VFD: ventilator free days; ARDS: acute respiratory distress syndrome.

In the UPMC-COVID cohort, from April 2020 through February 2022 we enrolled 49 patients with COVID-19 ARDS requiring IMV and obtained longitudinal plasma and ETA samples at baseline, middle and late intervals (Table S1). We performed 16S sequencing for bacteria and measured host response biomarkers in both sample types. In the MGH-COVID cohort, from April 2020 to May 2021we enrolled 97 hospitalized patients, obtained serial lung (sputum or ETA) and stool (gut) samples (Table S1) and performed Illumina metagenomics.^15^ To contextualize microbiota analyses from critically ill patients, we incorporated previously generated 16S-Seq data from upper respiratory tract (URT), LRT and stool samples collected from healthy volunteers (**Healthy Controls**), as previously described in smaller cross-sectional studies from our group.^11,12^

### Progressive dysbiosis of microbial communities in three body compartments

Among all three cohorts and healthy controls, we analyzed a total of 2557 clinical samples and 233 experimental control samples, with the latter obtained either during patient sampling at the bedside or during sample processing in the laboratory. In an initial quality control step, we demonstrated robust detection of bacterial 16S reads in oral, lung and gut samples in the UPMC-ARF cohort compared to negative controls (Figure S1A-B). We also found that rectal swabs not coated by stool (“unsoiled” swabs) had systematic differences in bacterial load (16S rRNA gene copies by qPCR) and beta diversity (Manhattan distances) compared to stool or visibly “soiled” rectal swabs (Figure S1C-D). Therefore, we excluded “unsoiled” rectal swabs from further analyses because they may not offer sufficient representation of gut microbiota.^11^

Samples from critically ill patients had significantly lower alpha diversity (Shannon index) in each compartment compared to corresponding healthy control samples. Alpha diversity further declined in all three body compartments across longitudinal samples (Figure 1A). Similarly, baseline ICU samples had markedly significant differences in beta diversity from healthy controls (Figure 1B). Taxonomic composition comparisons showed depletion of multiple commensal taxa in ICU samples, with significant enrichment for *Staphylococcus* in oral and lung samples, and *Anaerococcus* and *Staphylococcus* in gut samples (Figure 1C-D-E). Among ICU samples, bacterial load quantification by 16S qPCR confirmed that the LRT had significantly lower biomass compared to URT (oral) and gastrointestinal tract (Figure 1F).

**Figure 1.**
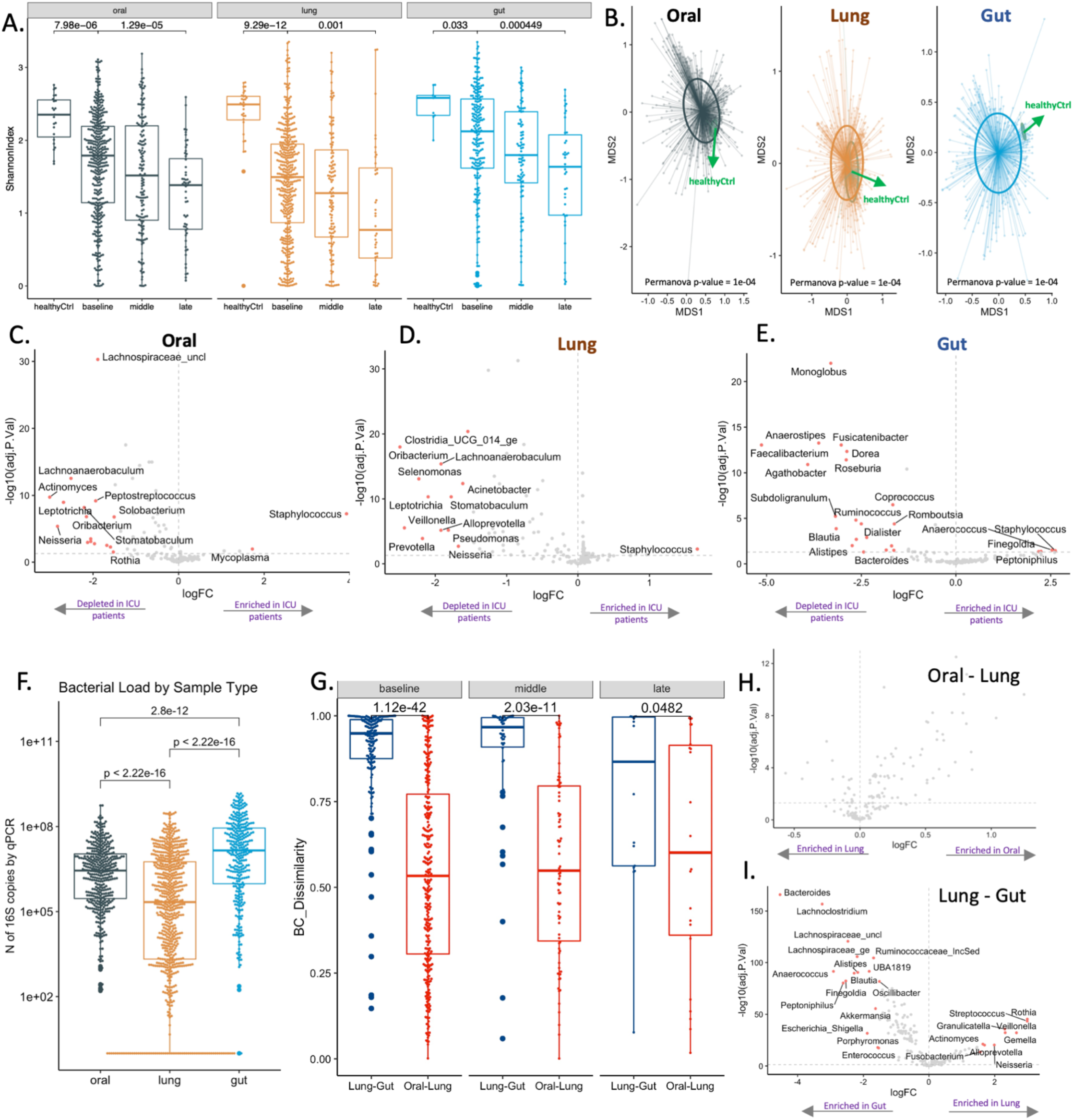
Ecological features of dysbiosis in three body compartments in critically ill patients. A. Samples from critically ill patients had significantly lower alpha diversity (Shannon index) compared to corresponding healthy control samples in each compartment (p<0.001), with further decline of Shannon index over time in longitudinal samples (p<0.001). B. Baseline samples from critically ill patients had markedly significant differences in beta diversity from healthy controls (permutational analysis of variance [permanova] p-values <0.001). C-E. Taxonomic composition comparisons with the *limma* package showed high effect sizes and significance thresholds (threshold of log2-fold-change [logFC] of centered-log-transformed [CLR] abundances >1.5; Benjamini-Hochberg adjusted p-value<0.05) showed depletion for multiple commensal taxa in critically ill patients samples, with significant enrichment for *Staphylococcus* in oral and lung samples, and *Anaerococcus* and *Enterococcus* in gut samples (significant taxa shown in red in the volcano plots). F. Lung samples had lower bacterial burden compared to oral and gut samples by 16S qPCR (all p<0.001). G. Oral and lung samples had higher compositional similarity (Bray-Curtis indices) compared to lung and gut samples in the baseline and middle interval (p<0.001). H-I: Taxonomic comparisons between compartments revealed that no specific taxa were systematically different between oral and lung microbiota (H), whereas in gut-lung comparisons, lung communities were enriched for typical respiratory commensals (e.g. *Rothia*, *Veillonella*, *Streptococcus*) and gut communities for gut commensals (e.g. *Bacteroides*, *Lachnoclostridium*, *Lachnospiraceae*) (I).

We then examined the compositional similarity (Bray-Curtis indices) between compartments to understand the relationship between the low biomass (lung) vs. high biomass (oral and gut) communities. We found higher similarity between oral-lung vs. gut-lung communities in the baseline and middle intervals (Figure 1G). Taxonomic comparisons between compartments revealed that no specific taxa were systematically different between oral and lung microbiota (Figure 1H), whereas in gut-lung comparisons, lung communities were enriched for typical respiratory commensals (e.g. *Rothia*, *Veillonella*, *Streptococcus*) and gut communities for gut commensals (e.g. *Bacteroides*, *Lachnoclostridium*, *Lachnospiraceae_uncl*) (Figure 1I). We specifically tested whether certain patients had enrichment for gut-origin bacteria in their oral or lung samples despite no overall enrichment of the lung compartment for gut bacteria. We found that 4.8% and 8.1% of oral and lung samples, respectively, had >30% relative abundance for gut-origin bacteria (Fisher’s test p=0.03, Figure S2A), with progressively increased enrichment over time (Fisher’s test = 0.02, Figure S2B) in lung samples.

Importantly, the gut-origin taxa enrichment in these lung samples could not be fully explained by oropharyngeal colonization with such taxa (Figure S2C). Taken together, these multi-site analyses point to the oral cavity as the primary source of lung microbiota, which could be seeded by micro-aspiration along the respiratory tract’s gravitational gradient. At the same time, our analyses also provided evidence for gut-origin bacteria enrichment in the LRT in a subset of critically ill patients.

We next examined the longitudinal composition of microbial communities by classifying bacteria in terms of their oxygen requirements (obligate anaerobes, facultative anaerobes, aerobes, microaerophiles, variable or unclassifiable) and plausible respiratory pathogenicity (oral commensals, recognized respiratory pathogens or other).^12^ In both oral and lung communities, we found a progressive decline in the relative abundance of obligate anaerobes over time. There was, however, no corresponding change in the gut composition of anaerobic (obligate or facultative) bacteria over time (Figure 2A-B). Stratified by plausible pathogenicity, we found a progressive decline of oral commensal bacteria in all three compartments, with a corresponding increase in pathogen abundance (Figure 2C-D). Fungal ITS sequencing showed that >50% of communities in all three compartments were dominated by *C. albicans* (defined as >50% relative abundance), with a progressive decline in fungal Shannon index in oral and lung communities during follow-up (Figure S3). Nanopore metagenomics of lung samples provided similar bacterial representations to 16S analyses and confirmed high abundance of *C.albicans* detected by ITS sequencing (Figure S3). Thus, our analyses revealed a pattern of compartment-wide dysbiosis in ICU patients, with progressive decline in diversity and enrichment for plausible pathogenic bacteria and *C. albicans*. We then sought to understand whether patient-level variables accounted for baseline or longitudinal dysbiosis.

**Figure 2:**
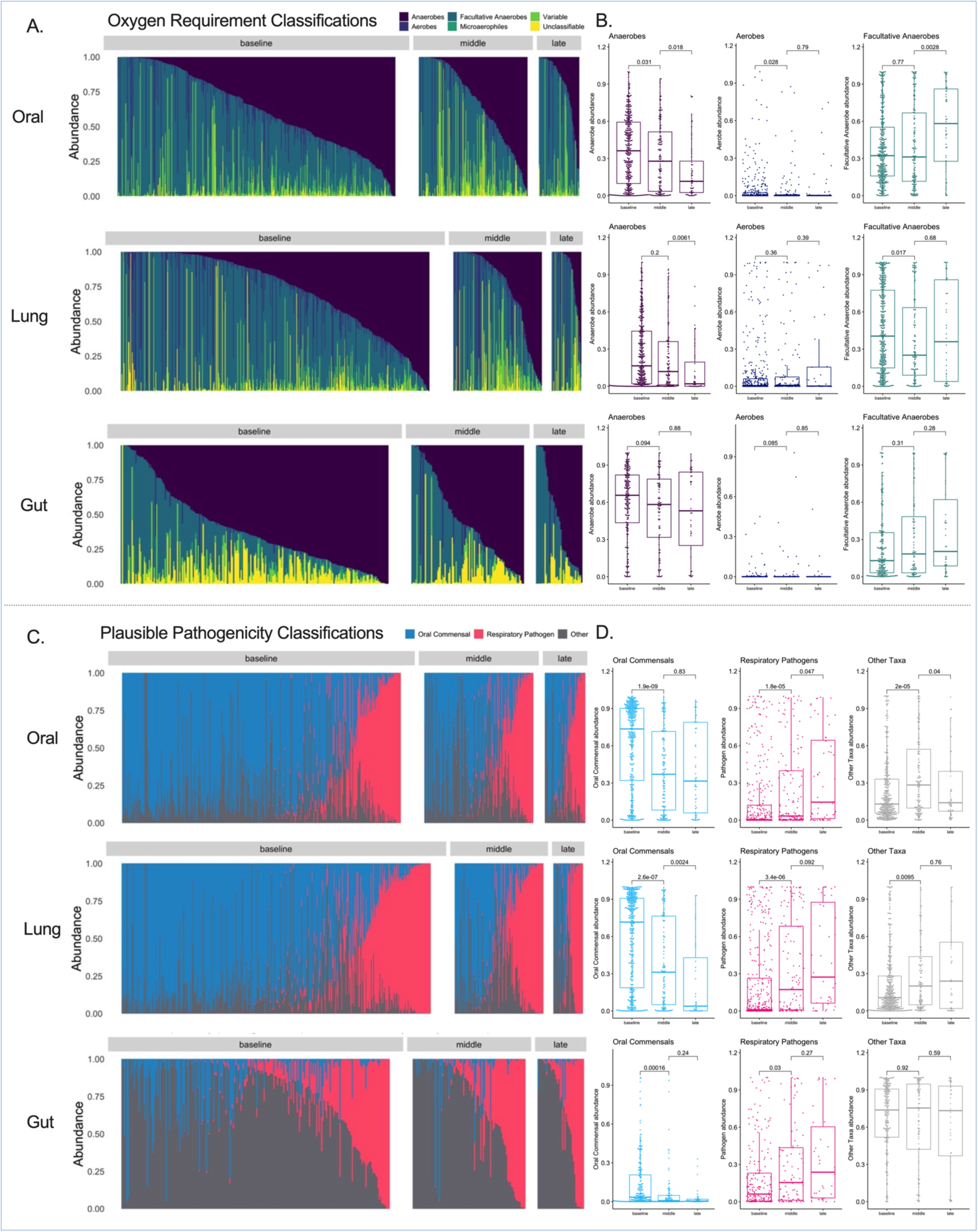
Longitudinal analysis of bacterial composition showed a progressive loss of obligate anaerobes in oral and lung communities as well as enrichment for recognized respiratory pathogens in all three compartments. Top Panels (A-B): Relative abundance barplots for oral, lung and gut samples with classification of bacterial genera by oxygen requirement into obligate anaerobes (anaerobes), aerobes, facultative anaerobes, microaerophiles, genera of variable oxygen requirement and unclassifiable. Comparisons of relative abundance for the three main categories of bacteria (obligate anaerobes, aerobes and facultative anaerobes) by follow-up interval (baseline, middle and late). Data in boxplots (B) are represented as individual values with median values and interquartile range depicted by the boxplots with comparisons between intervals by non-parametric tests. Bottom Panels (C-D): Relative abundance barplots for oral, lung and gut (F) samples with classification of bacterial genera by plausible pathogenicity into oral commensals, recognized respiratory pathogens and “other” category. Comparisons of relative abundance for these categories of bacteria by follow-up interval (baseline, middle and late) in boxplots (D).

### Clinical diagnoses and antibiotic exposure correlate with microbial community diversity and composition

We constructed linear regression models with ecological metrics indicative of dysbiosis as outcomes (baseline Shannon index, obligate anaerobe and respiratory pathogen abundance) and clinical variables as predictors (Figure S4). History of COPD, immunosuppression and clinical diagnosis of pneumonia showed the most significant associations with dysbiosis features, e.g. lower Shannon and anaerobe abundance in oral and lung communities for patients with COPD, and increased pathogen abundance in all three compartments for patients with history of immunosuppression (Figure S4). History of immunosuppression was also associated with higher abundance of *C. albicans* in oral and lung samples (Figure S3D). To further explore iatrogenic forces on microbiota composition, we focused on two common treatments in the ICU: antibiotics and steroids. We assessed antibiotic usage by i) anaerobic coverage, ii) a numerical scale that included duration, timing and type,^16^ and iii) the Narrow Antibiotic Treatment (NAT) score.^12,17^ We quantified steroid use as the daily equivalent dosage of prednisone in milligrams. Antibiotic usage was associated with Shannon index, anaerobe, and pathogen abundance in baseline gut samples, with exposure to antibiotics with anaerobic spectrum at baseline being inversely correlated with anaerobe abundance in all three compartments (Figure S4B). To explore the effects of antibiotics and steroids over time, we employed mixed linear regression models using longitudinal samples. In all three compartments, the receipt of anaerobic spectrum antibiotics was associated with a progressive decrease in obligate anaerobe abundance, without significant effects on pathogen abundance (Table S2). Notably, antibiotic exposure quantified by the NAT score was also significantly linked to a reduction in anaerobe abundance and an increase in pathogen abundance within the gut microbiota. Steroids were associated with decrease in anaerobes in the lungs, but not with changes in abundance of other microbes in other compartments.

### Microbial communities in each compartment form distinct clusters of diversity and composition

We next examined the microbial communities independent of clinical variables to capture important features directly from microbiome data. To understand microbial heterogeneity within compartments, we leveraged two complementary unsupervised clustering approaches: i) Dirichlet Multinomial Mixture (DMM) models for 16S data in each compartment (“bacterial DMM clusters”) and for Nanopore metagenomic data in the lung compartment^18^, and ii) weighted Similarity Network Fusion (SNF)^19^ clusters for combined bacterial (16S) and fungal (ITS) data within each compartment (“bacterial-fungal SNF clusters”).

By bacterial DMM clusters, a three-class model offered optimal classification in each compartment, with striking differences in alpha diversity and composition between clusters (Figure 3A). Cluster 1 in each compartment had high Shannon index in the range of healthy controls (referred to as High-Diversity cluster), cluster 3 had low Shannon index (Low-Diversity cluster), and cluster 2 had intermediate diversity (Intermediate-Diversity cluster). Low-diversity clusters had markedly higher abundance of pathogens and lower abundance of anaerobes (Figure 3B-C). In cross-compartment comparisons, DMM cluster membership was strongly associated between oral and lung communities (odds ratio of membership in the Low-Diversity cluster in both compartments 9.74, 95% confidence interval [5.61-17.29], p<0.0001), whereas lung and gut clusters were less strongly associated although statistically significant (p=0.015, Figure 3D). In longitudinal analyses, cluster membership showed relative stability for all compartments, with most samples assigned to Low-Diversity cluster at baseline being assigned to Low-Diversity in the middle interval as well (77% of oral, 80% of lung, and 78% of gut samples, respectively, Figure S5). Nanopore DMM clustering in 130 available lung samples also showed optimal fit with three total clusters (data not shown). Bacterial-Fungal SNF clustering revealed distinct communities in each compartment, with a notable cluster in lung samples (cluster 1) with high pathogen abundance and *C. albicans* dominance (near 100% of fungal sequences abundance) (Figure S6). Thus, our unsupervised clustering approaches captured broad differences in meta-communities that were not specific to individual taxa. We next examined how these microbial communities related to host responses and clinical outcomes.

**Figure 3:**
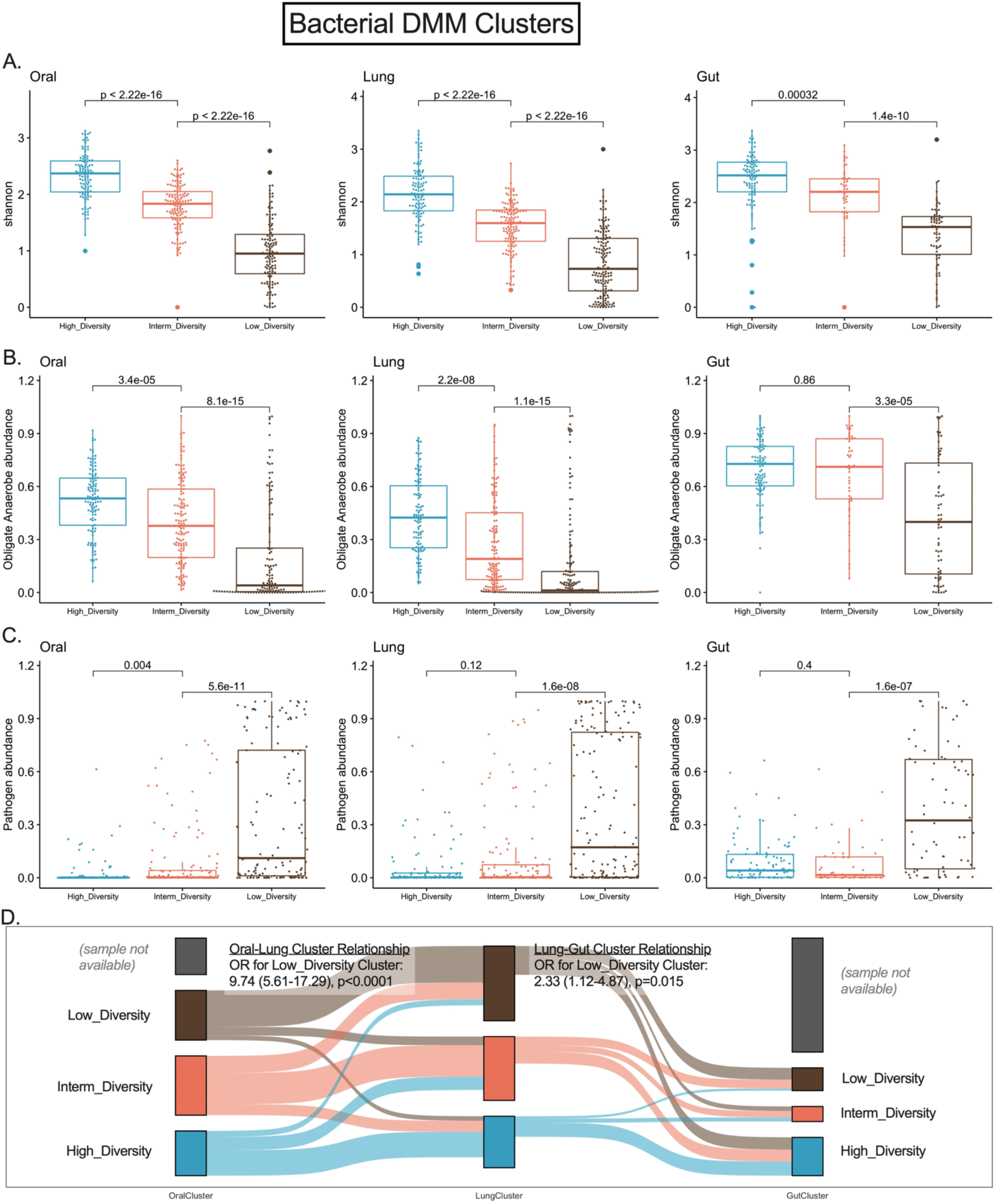
Unsupervised clustering approaches revealed differences in bacterial alpha diversity and composition in three body compartments of critically ill patients. Panels A-D demonstrate bacterial Dirichlet Multinomial Mixture (DMM) modeling results for each compartment separately. DMM clusters had significant differences in alpha diversity (A) and composition (obligate anaerobe abundance in shown in panel B and pathogen abundance shown in panel C), with cluster 3 in each compartment showing very low Shannon Index and enrichment for pathogens (Low-Diversity cluster). Oral and lung cluster assignments were strongly associated (Odds ratio for assignment to the Low-Diversity cluster: 9.74 (5.61-17.29), p<0.0001), whereas lung and gut cluster assignments were less strongly but significantly associated (panel D).

### Lung microbiota correlate with systemic host responses

We examined host-microbiota interactions with two independent approaches, a microbiota– and a host-centric approach. In the microbiota-centric approach, we correlated the top 20 abundant taxa in each compartment with systemic (plasma) and lung-specific (ETA/BALF supernatants) host response biomarkers. We found several significant correlations (Figure S7A-C), with typical pathogens (e.g. *Klebsiella*, *Escherichia-Shigella*, *Staphylococcus* genera in the lung compartment) positively correlating with plasma inflammatory biomarkers (such as sTNFR1 and IL-6 levels), whereas typical oral commensals (e.g. *Rothia*, *Streptococcus*, *Prevotella* etc.) inversely correlated with plasma sTNFR1 or sRAGE. In cluster comparisons, the bacterial DMM Low-Diversity cluster in the lungs was significantly associated with higher plasma sTNFR1, sRAGE and procalcitonin levels (Figure S7D), whereas the Nanopore DMM Low-Diversity cluster was also significantly associated with higher regional (IL-6 and sRAGE) and systemic biomarkers of injury and inflammation (plasma IL-6, sTNFR1, sRAGE, Ang-2 and Pentraxin-3, Figure S7E).

In the host-centric approach, we applied a widely validated framework of host-response subphenotypes based on plasma biomarkers.^7,20^ With a validated 4-biomarker parsimonious model (using sTNFR1, Ang2, procalcitonin and bicarbonate levels),^20^ we classified individuals at baseline into a hyperinflammatory (22.9%) vs. a hypoinflammatory (77.1%) subphenotype. We found no significant relationship between host subphenotypes and DMM microbiota clusters in any compartment (Figure S7G), but hyperinflammatory patients had higher pathogen abundance in lung communities (p=0.04). To further investigate this association, we stratified patients by pneumonia diagnosis. We discovered that hyperinflammatory patients without pneumonia had higher pathogen abundance in lung samples compared to hypo-inflammatory patients (p=0.018, Figure S7H). These notable associations between lung pathogen abundance and the hyperinflammatory subphenotype imply that systemic subphenotypes might stem, at least in part, from undiagnosed pneumonia or respiratory dysbiosis.

### Lung microbiota clusters predict survival independent of clinical variables and host responses

Comparisons of microbial communities between survivors and non-survivors at 60-days post-ICU admission showed highly significant differences in alpha diversity in the lungs (p<0.0001), as well as higher obligate anaerobe and lower pathogen abundance in both oral and lung samples (all p<0.002, Figure S8A-C), but no differences in gut profiles. Additionally, analyses of lung samples stratified by whether they exhibited gut-origin taxa enrichment (defined as >30% relative abundance) showed markedly worse survival for patients with gut-origin taxa enrichment (p<0.0001, Figure S2E-F).

Analyses by bacterial DMM clusters provided further insights with regards to the prognostic value of each compartment. In both oral and lung compartments, the Low-Diversity clusters were associated with worse 60-day survival in Kaplan-Meier curve analyses, whereas gut clusters had no survival impact (Figure 4A-C).

**Figure 4:**
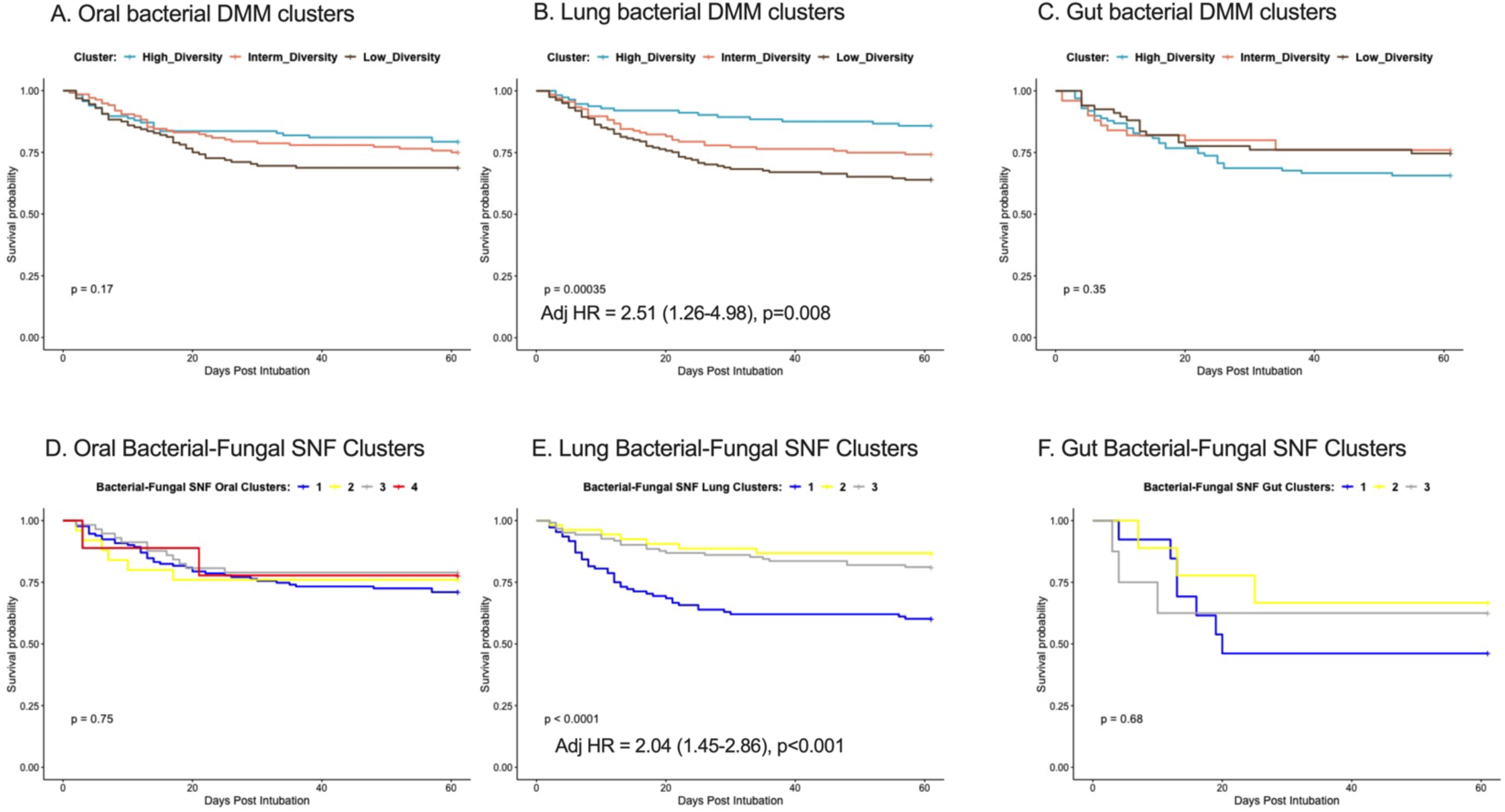
Lung bacterial and bacterial-fungal clusters strongly predicted 60-day survival independent of clinical predictors, organ dysfunction severity and host-response subphenotypes. A-C: Kaplan-Meier curves for 60-day survival from intubation stratified by oral (A), lung (B) and gut (C) bacterial DMM clusters. The Low-Diversity lung DMM cluster was independently predictive of worse survival (adjusted Hazard Ratio = 2.51 (1.26-4.98), p=0.008), following adjustment for age, sex, history of COPD, immunosuppression, severity of illness by sequential organ failure assessment (SOFA) scores and host-response subphenotypes. The Lung bacterial-fungal SNF cluster with high pathogen and C. albicans abundance (cluster 1) was independently predictive of worse survival (D), whereas the oral and gut bacterial-fungal SNF clusters (D, F) did not impact survival.

Notably, the prognostic effects of the Low-Diversity bacterial DMM cluster in the lungs remained significant after adjustment for age, sex, history of COPD, immunosuppression, severity of illness by SOFA scores and host-response subphenotypes (adjusted Hazards Ratio-HR= 2.51 [1.26-4.98], p=0.008). Similarly, survival analysis by the bacterial-fungal SNF lung clusters showed that cluster 1, which had high pathogen and *C. albicans* abundance, also independently predicted worse survival (adjusted HR=2.04 [1.45-2.86], p<0.0001, Figure 4E). The other bacterial-fungal SNF oral and gut clusters did not impact survival (Figure 4D,F). Thus, we found evidence that lung microbiota dysbiosis predicted survival beyond the information provided by clinical predictors, commonly used organ dysfunction indices, and biological subphenotyping.

### Derivation of a dysbiosis index and external validation in patients with COVID-19

Motivated by the robust, independent prognostic impact of microbiota clusters on patient survival, we next sought to construct predictive models to classify bacterial profiles into the corresponding DMM clusters within each compartment. Such predictive models could serve as dysbiosis indices beyond the derivation cohort with our DMM analysis. We used probabilistic graphical modeling (PGM) to predict the DMM clusters in each compartment based on the abundance of the top 50 taxa and the corresponding Shannon index. By splitting the dataset in training and testing subsets (80% and 20% of data points, respectively), we developed separate multinomial regression models for DMM cluster predictions in each compartment (i.e. compartment-specific Dysbiosis Index), which showed accuracy of 0.76, 0.86 and 0.75 for oral, lung and gut clusters, respectively. We verified that patients classified in the low diversity clusters by the Dysbiosis Index for the oral and lung compartments had worse survival, similarly to the DMM-derived clusters.

We next applied the derived Dysbiosis Indices to two independent cohorts of hospitalized patients with COVID-19 pneumonia. In the UPMC-COVID cohort of patients with COVID-19 ARDS on IMV (n=49), the Lung Dysbiosis Index classified ETA samples into three clusters with significant differences in Shannon index and bacterial load by qPCR (Figure 5A), but no difference in ETA SARS-CoV-2 viral load by qPCR or 60-day survival (data not shown). Patients assigned to the low diversity cluster at baseline had higher plasma levels of sTNFR1 and Ang-2 compared to the high diversity cluster (p<0.05, Figure 5B). By individual taxa abundance, oral commensals (e.g. *Prevotella*, *Veillonella* or *Streptococcus*) were inversely correlated with plasma sTNFR1 and Ang-2, whereas *Klebsiella* abundance was positively correlated (all p<0.05), corroborating the findings of the cluster analyses relating lung microbiota with prognostically adverse higher levels of systemic biomarkers of inflammation and endothelial injury.

**Figure 5:**
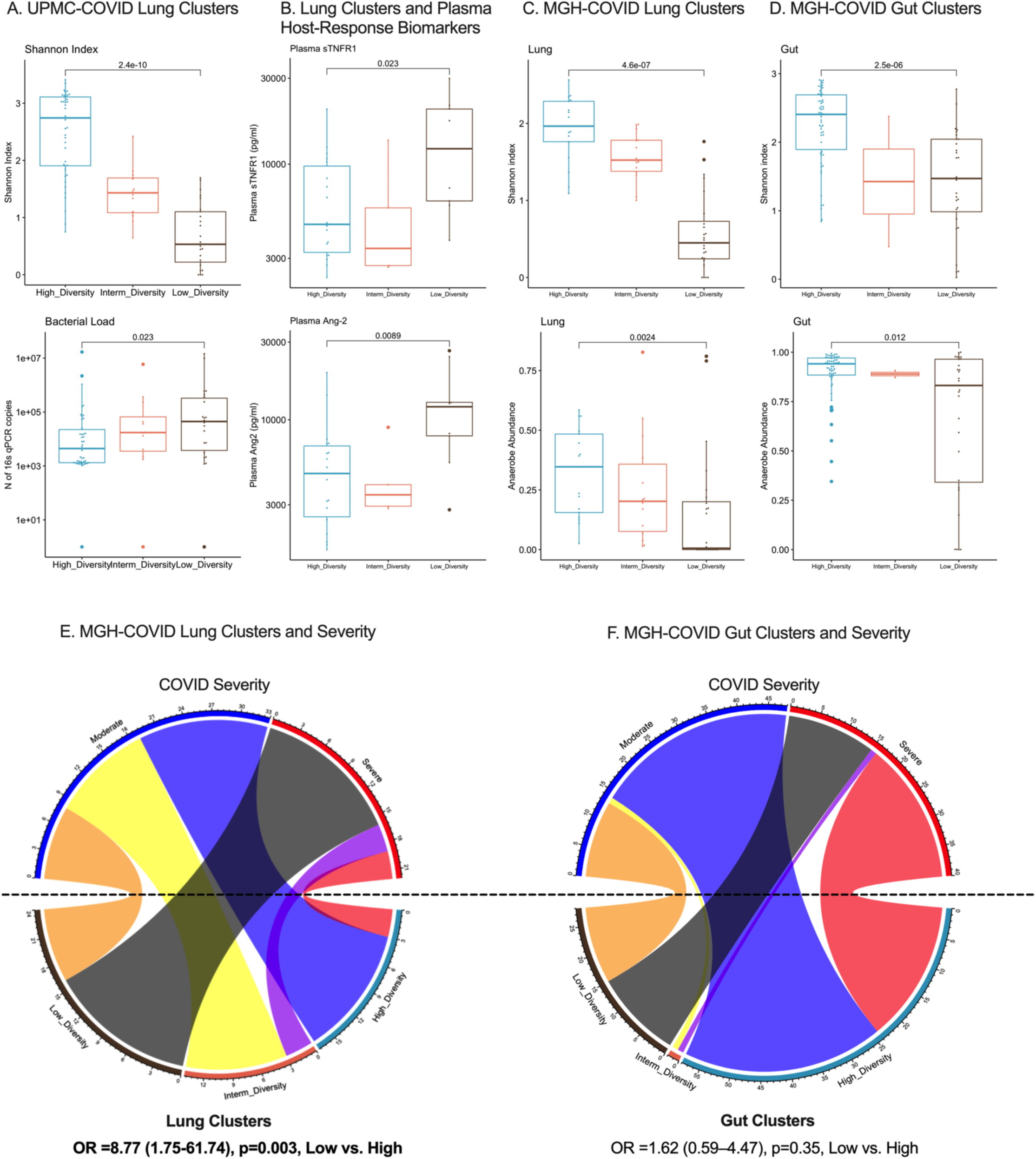
**Lung and Gut Microbiota Associations with COVID-19 Severity in Two Independent Cohorts**. A. Application of the dysbiosis index in lung (ETA) microbiota profiles in the UPMC-COVID cohort classified subjects in three clusters, with significant differences in Shannon index and bacterial load by 16S qPCR. B. The low diversity cluster in lung samples from UPMC-COVID subjects was significantly associated with higher plasma levels of sTNFR1 and Ang-2. C-D. Application of the dysbiosis index models in lung (sputum or ETA) and gut (stool) samples in the MGH-COVID cohort classified subjects in three clusters, with significant differences in Shannon index and anaerobe abundance between clusters. E-F: Cluster assignments in the MGH cohort were strongly associated with clinical severity for lung samples only. Membership in the Low-Diversity cluster in the lungs was associated with an odds ratio of 8.77 (1.75-61.74) for severe disease (black belt connecting the Low-Diversity cluster and Severe Disease perimetric zones in the chord diagram). Gut clusters were not significantly associated with clinical severity of COVID-19 pneumonia.

In the MGH-COVID cohort (n=97), we performed metagenomic sequencing in longitudinal lung (ETA for patients on IMV or expectorated sputum in spontaneously breathing patients) and gut (stool) samples obtained upon enrollment and then daily up to day 4. We found no significant changes over time in Shannon Index and anaerobe/pathogen abundance in either compartment on serial samples through day 4. We classified baseline lung and gut samples by our Dysbiosis Index models, which showed significant differences in Shannon index, anaerobe and pathogen abundance in each compartment (Figure 5C-D). Importantly, the low diversity cluster in the lung compartment was strongly associated with COVID-19 pneumonia severity (odds ratio 8.77 [1.75-67.74], Figure 5E-F), as classified by oxygen support requirements, whereas gut clusters were not. Thus, application of the Dysbiosis Indices to lung and gut samples of patients with COVID-19 provided similar findings to the ones obtained in the UPMC-ARF derivation cohort, supporting the predictive value of lung microbiota profiling.

## Discussion

We conducted a longitudinal, integrative assessment of host-microbiota interactions in a large cohort of ARF patients across three body sites (the oral cavity, lungs, and gut) and up to three time-points in the ICU. These analyses offered insights into the temporal relationships between patient-level factors, therapeutic interventions, microbial communities and patient-centered outcomes, which has not been possible in previous smaller scale investigations.^21^ The progressive dysbiosis of microbial communities observed in all three body compartments highlights the impact of critical illness on the global microbiota. We found reduced alpha diversity and deviation in composition compared to healthy controls at the onset of IMV, with further reduction in diversity and alterations in composition for patients supported on ventilators over time. Unsupervised analyses of microbiota composition revealed distinct communities in all three body compartments, yet the lung microbiome emerged as the strongest independent predictor of important clinical outcomes. We developed parsimonious models for dysbiosis classifications in each compartment and found that lung dysbiosis was significantly associated with host-response profiles and clinical severity in patients with COVID-19.

The large sample size and granular clinical data in our derivation cohort allowed for detailed investigation of the relationships between patient-/treatment-related factors with the composition of microbiota. Clinical diagnoses (e.g., ARDS or pneumonia) and comorbidities explained variation in diversity and composition at baseline. We detected significant associations between systemic steroid exposure and lung microbiota composition, a novel finding that warrants validation in other cohorts. Despite the self-evident biological plausibility of antibiotic pressures on altering the microbiomes of critically ill patients, empirical evidence to date has been limited.^22–24^ Here we modeled antibiotic exposure thoroughly with different methodologies from prior studies focused on cystic fibrosis or pneumonia,^16,17,25^ and studied antibiotic effects on longitudinal communities and features of dysbiosis. We found that the NAT score and a simple categorical classification with regards to anaerobic spectrum coverage captured important effects on longitudinal composition. Recent epidemiologic and molecular evidence supports disruptive effects of anti-anaerobic antibiotics in gut microbial communities.^24,26^ Our data are consistent with the idea that anaerobe-targeting antibiotics are associated with anaerobic bacteria depletion in the respiratory and the intestinal tracts, and furthermore our study suggests that such depletion is associated with worse clinical outcome. Therefore, our results highlight the importance of rational use of anti-anaerobic antibiotics as directed by proper clinical indications, because such antibiotics can have important yet under-recognized adverse clinical implications.

The biogeography of the intubated respiratory tract has been the focus of extensive investigation for prevention of secondary ventilator-associated pneumonia (VAP).^27,28^ Oropharyngeal decontamination with chlorhexidine rinses or the more aggressive selective digestive decontamination (SDD) of the gastrointestinal tract have been studied for reducing bacterial burden in probable source compartments that seed the LRT microbiota. While both decontamination approaches are supported by randomized clinical trial evidence showing efficacy in VAP prevention^29,30^, both approaches also have associated safety concerns,^31,32^ leading tto limited uptake of SDD worldwide. Indiscriminate application of chlorhexidine rinses in all patients on IMV may also deplete commensal organisms from the URT and reduce colonization resistance against pathogens. We found significant correlations between oral-origin commensal taxa abundance in URT and LRT samples, such as *Prevotella*, with prognostically favorable, lower levels of plasma inflammatory biomarkers, which may indicate favorable regulation of innate immunity by such taxa.^33–35^ Our comparative analyses between compartments showed much higher oral-lung than lung-gut similarity, suggesting that the oral cavity serves as the primary source of microbial seeding for the lungs. However, we found that a small subset of patients had enrichment for gut-origin commensal or pathogenic organisms in their LRT, which could not be fully accounted for by URT colonization with similar taxa. Such patients with gut-origin bacteria enrichment in their lungs (8.1%) had much worse survival than the rest of the cohort, and may represent a subset of patients in whom gut-to-lung bacterial translocation may have occurred.^36,37^ Wider availability of BAL samples to investigate the alveolar spaces more closely can provide more evidence into the question of gut-to-lung translocation, but our non-invasive ETA samples showed that such translocation, if present, affects a small subset of patients at least within the first week of IMV. Therefore, efforts focused on preventing dysbiosis and pathogen colonization in the URT-to-LRT ecosystem may offer higher biological plausibility for measurable benefits in clinical trials.

Unsupervised clustering revealed distinct microbial communities within and across body compartments. Low-diversity bacterial clusters were enriched with pathogens and depleted in anaerobes in all three compartments. Membership in the low-diversity cluster was strongly associated between the oral and lung compartment, suggesting shared patterns of dysbiosis. The overall stability of longitudinal cluster membership indicated that specific microbial profiles may persist throughout critical illness, influencing the disease trajectory. Integration of fungal sequencing data further enhanced our view of the microbial communities, revealing patients who had a “double-hit” of bacterial pathogen enrichment and *C.albicans* dominance in their communities. We have recently shown that *C.albicans* abundance in the LRT correlates with systemic inflammation and predicts adverse outcome in patients with ARF on IMV.^13^ With the current expanded dataset, we demonstrate that integration of bacterial and fungal data can identify patient subpopulations with inter-kingdom dysbiosis, who may require different interventions to address both bacterial and fungal dysbiosis.

Survival analyses based on microbiota clusters revealed two significant and novel findings. First, in this comparison of microbiota from three distinct body compartment microbiota for predicting survival in critically ill patients, the lung microbiome emerged as the most powerful predictor compared to oral or gut microbiota. Perhaps this finding should not be surprising when studying patients who required IMV for ARF. We had previously shown that baseline lung microbiota profiles were predictive of survival.^3^ We now expand analyses to three compartments up to three time points during IMV and show that lung microbiota carry the most predictive signal for survival, both at baseline and also in follow-up samples. Thus, our comparative assessment of microbiota across body compartments highlights the clinical relevance of lung microbiota analysis in critical illness and the need for dedicated sampling of the LRT.^38^ Second, the prognostic value of lung microbiota clusters was independent not only from clinical predictors and validated organ dysfunction metrics, such as the SOFA score, but also from the systemic host-response subphenotypes. Extensive evidence has established the prognostic value and generalizability of plasma biomarker-based subphenotyping of patients with ARF.^8,39^ Our adjusted Cox proportional hazards models revealed significant hazards ratios for the Low-Diversity lung cluster, when analyzed using both the bacterial DMM and bacterial-fungal SNF methods. Beyond the significant taxa-biomarker associations we observed, the survival analyses demonstrated that lung microbiota may influence patient outcome in ways that are not captured by current host-response subphenotyping approaches. An integrative, host– and lung microbiome-aware subphenotyping framework may thus augment our ability to better prognosticate and target therapeutic interventions in ARF.

Our study has several limitations. First, we mainly focused on bacterial and fungal components of the microbiome, and thus could not assess the role of the virome, especially with regards to respiratory RNA viruses. The consistent pattern of results relating elements of the bacterial microbiome to host response and illness severity in the COVID-19 cohorts supports the generalizability of our findings, although we could not investigate contributions from individual viruses. The observational nature of our study prevents us from establishing causality between the microbiome and clinical outcomes, which could be addressed by future interventional studies or animal modeling with microbiome manipulation. Longitudinal sample availability was limited by informative censoring, as patients with rapid decline and early death or those with rapid improvement and liberation from IMV would not contribute follow-up samples in the middle and late intervals. We aimed to mitigate some of these right censoring biases with mixed linear regression models, but our longitudinal analysis findings should be interpreted with caution and considered as applicable to patients who remain on IMV for the first 1-2 weeks of critical illness. For patient safety and practical purposes of subject participation in our observational research study, we relied on non-invasive biospecimens (ETA) for LRT microbiota profiling, as opposed to reference standard BAL.^38^ Our non-invasive approach allowed us to enroll a large cohort of LRT specimens, follow serial samples over time, and is congruent with clinical practice guidelines for VAP diagnosis.^40^ However, we may have missed important microbiota variability closer to the alveolar space, including a stronger signal of gut-to-lung microbiota translocation.^37^ Finally, we had a smaller effective sample size for gut microbiota analysis, which may have limited our ability to identify prognostic variation within the gut compartment.

In conclusion, our study provides novel insights into the predictive value of microbiota clusters derived from different body compartments in critically ill patients. The lung microbiome emerged as the most powerful predictor of survival, surpassing the oral and gut microbiota. These findings emphasize the clinical relevance of investigating the lung microbiota and highlight its potential as a prognostic marker in critical illness. Moreover, our study underscores the importance of considering organ-specific microbial communities in critical care settings and expands our understanding of the microbiome’s role in determining patient outcomes. Further research in this area has the potential to shape clinical decision-making and facilitate the development of personalized medicine strategies for critically ill patients.

## Online Methods

### UPMC-ARF cohort

Following admission to the ICU at UPMC (Pittsburgh, PA, USA) and obtaining informed consent from patients or their legally authorized representatives (University of Pittsburgh IRB protocol STUDY19050099), we collected baseline research biospecimens within 72hrs from intubation. We collected blood for separation of plasma, oropharyngeal (oral) swabs to profile upper respiratory tract (URT) microbiota, endotracheal aspirates (ETA) for LRT (lung) microbiota and host biomarker measurements, and rectal swabs or stool samples for gut microbiota analyses. We also captured leftover bronchoalveolar lavage fluid (BALF) from clinically indicated bronchoscopies, when available. We repeated research biospecimen sampling between days 3-6 (middle interval) and days 7-12 (late interval) post enrollment for subjects who remained in the ICU. No patients in the UPMC-ARF cohort were known to be infected by SARS-CoV-2 at the time of enrollment.

### UPMC-COVID cohort

Following admission to the ICU and obtaining informed consent from patients or their legally authorized representatives (University of Pittsburgh IRB protocol STUDY19050099), we collected baseline research biospecimens (ETA and blood) within 72hrs from intubation. We repeated research biospecimen sampling between days 3-6 (middle interval) and days 7-12 (late interval) post enrollment for subjects who remained in the ICU, as per the UPMC-ARF protocol. All patients were known to be infected by positive SARS-CoV-2 qPCR prior to enrollment.

### MGH-COVID cohort

From April 2020 to May 2021, we prospectively enrolled 97 hospitalized patients aged ≥18 years with confirmed COVID-19 at the Massachusetts General Hospital (Boston, MA, USA) to a longitudinal COVID-19 disease surveillance study.^15^ The Study protocol #2020P000804 was approved by the Mass General Brigham IRB. All participants or their healthcare proxy provided written informed consent to participate. Patients were categorized as having severe COVID-19 if they required admission to the intensive care unit with acute respiratory failure (the need for oxygen supplementation ≥15 liters per minute (LPM), non-invasive positive pressure ventilation, or mechanical ventilation) or other organ failure (such as shock requiring vasopressors). Otherwise, they were categorized as having moderate COVID-19. Expectorated sputum, ETA or fresh stool was collected and refrigerated at 4℃ until aliquoting/freezing at –80℃ (typically within 4 hours of collection) from adult patients enrolled in the prospective biospecimen collection study. Participants were able to provide samples as frequently as once daily for up to four days, as well as declining donation on any given day (while remaining in the study).

### Healthy Controls

To contextualize the findings on microbiota from critically-ill patients with what is expected for the healthy respiratory and gastrointestinal tract, we also included data from 24 healthy volunteers who had contributed URT and LRT microbiome data in a previously published cohort (Lung HIV Microbiome Project – University of Pittsburgh IRB STUDY19060243),^41^ as well as stool from 15 healthy donors for fecal microbiota transplantation (University of Pittsburgh IRB – STUDY20060312).^11^ We designated these healthy volunteers as Healthy Controls.

### Clinical data recording

A consensus committee reviewed clinical and radiographic data and performed retrospective classifications of the etiology and severity of acute respiratory failure without knowledge of microbiome sequencing or biomarker data. We retrospectively classified subjects as having ARDS per established criteria (Berlin definition), being at risk for ARDS because of the presence of direct (pneumonia or aspiration) or indirect (e.g., extrapulmonary sepsis or acute pancreatitis) lung-injury risk factors although lacking ARDS diagnostic criteria, having acute respiratory failure without risk factors for ARDS, or having acute-on-chronic respiratory failure. We followed patients prospectively for cumulative mortality and ventilator-free days (VFDs) at 30 days, as well as survival up to 60 days from intubation.

We systematically reviewed administered antibiotic therapies since hospital admission and recorded the antibiotic exposure for each subject according to the following three metrics:

1. Anaerobic coverage (yes/no): whether antibiotics with anaerobic coverage were given on the day of sampling.
2. The Antibiotic Exposure score by Zhao et al ^16^: a numerical scale with antibiotic weighting based on dosing duration, timing of administration relative to sample collection and antibiotic type and route of administration. We utilized the convex increasing weighting scheme and modeled the antibiotic exposure from hospital admission until day of sampling.
3. The Narrow Antibiotic Treatment (NAT) score developed for community-acquired pneumonia treatment studies ^17,25^. We calculated the daily NAT score from –5 days from sampling to post 10 days after sampling on day 1.

### Research Sample Collection

Within the first 48 hours of intubation (baseline time-point), we collected a posterior oropharyngeal (oral) swab via gentle swabbing the posterior oropharynx next to the endotracheal tube with a cotton tip swab for 5 secs, and an endotracheal aspirate (ETA) via suctioning secretions from the endotracheal tube with the in-line suction catheter and without breaking seal in the ventilatory circuit.^1,4^ Rectal swabs were collected according to a standard operating procedure (i.e., placing the patient in a lateral position, inserting the cotton tip of the swab into the rectal canal, and rotating the swab gently for 5 s), unless clinical reasons precluded movement of the patient (e.g., severe hemodynamic or respiratory instability). Stool samples were collected when available, either by taking a small sample from an expelled bowel movement (before cleaning of the patient and disposal of the stool) or from a fecal management system (rectal tube) placed for management of diarrhea and liquid stool collection. We also collected simultaneous blood samples for centrifugation and separation of plasma. Samples were delivered to the processing laboratory within minutes from acquisition, and then aliquoted and stored in –80C until conduct of experiments. For samples that underwent host DNA depletion for Nanopore sequencing, an aliquot remained in 4C for processing before freezing for up to 72hrs from acquisition. ETA Samples obtained from COVID-19 subjects inactivated by 4-fold dilution in DNA/RNA Shield (Zymo Research) under biosafety level 2+ conditions and then stored at −80°C. For patients who remained intubated in the ICU, we collected follow-up samples at a middle time-point (days 3-6) and a late follow-up interval (days 7-11 post-intubation).

For Healthy Controls, an oral wash and BAL sample were collected with a standardized protocol.^41^ Subjects were asked to fast and refrain from smoking for at least 12hrs before sample collection. Oral washes were performed by having participants gargle with 10 ml sterile 0.9% saline immediately before bronchoscopy. BAL was performed according to standardized procedures developed to minimize oral contamination.

Participants gargled with an antiseptic mouthwash (Listerine) immediately before topical anesthesia. The bronchoscope was then inserted through the mouth and advanced to a wedge position quickly and without use of suction. BAL was performed in the right middle lobe or lingula up to a maximum of 300 ml 0.9% saline.

Healthy donors of stool for fecal microbiota transplant collected a stool sample in a specialized container and brought the stool sample on the day of collection to the processing lab.

### Laboratory Analyses

#### Microbiome assays in UPMC cohorts

From all available oral swabs, ETAs, left over BALF, rectal swabs and stool samples, we extracted genomic DNA and performed quantitative PCR (qPCR) of the V3-V4 region of the 16S rRNA gene to obtain the number of gene copies per sample, as a surrogate for bacterial load. From a separate aliquot of extracted DNA from oral swabs, ETA, rectal swabs and stool samples, we performed amplicon sequencing for bacterial DNA (16S-Seq of the V4 hypervariable region) and fungal DNA (ITS) on the Illumina MiSeq platform at University of Pittsburgh Center for Medicine and the Microbiome laboratories. ^3,42^ We used extensive experimental negative controls in all processing steps to rule out contamination, as well as mock microbial community positive controls (Zymo) to ensure target amplification success. We processed derived 16S sequences with a custom Mothur-based pipeline and performed analyses at genus level. From a random subset of 130 available ETA samples, we performed metagenomic Nanopore sequencing (following human DNA depletion) with a rapid PCR barcoding kit (SQK-RPB004) on the MinION device (Oxford Nanopore Technologies-ONT, Oxford, UK) for five hours.^43,44^ We analyzed microbial metagenomic sequences with the EPI2ME platform (ONT) and the “What’s In My Pot” [WIMP] workflow to quantify abundance of microbial species.^45^ We filtered FASTQ files with a mean quality (q-score) below a minimum threshold of 7.

#### Host-response assays

We measured 10 plasma biomarkers of tissue injury and inflammation with custom Luminex multi-analyte panels from plasma samples and ETA supernatants, when available. Specifically, we used a 10-plex Luminex panel (R&D Systems, Minneapolis, MI, United States) to measure interleukin(IL)-6, IL-8, IL-10, soluble tumor necrosis factor receptor 1 (sTNFR1), suppressor of tumorigenicity-2 (ST2), fractalkine, soluble receptor of advanced glycation end-products (sRAGE), angiopoietin-2, procalcitonin and pentraxin-3.^7^

#### Microbiome assays in MGH-COVID cohort

Samples were extracted and sequenced at Baylor College of Medicine according to their standard established platforms. DNA was prepared for sequencing using the Illumina Nextera XT DNA library preparation kit. All libraries were sequenced with a target of 3GB output at 2×150bp read length using the Illumina NovaSeq platform, as previously described.^15^

#### Quantification and statistical analysis

We performed non-parametric comparisons for continuous (described as median and interquartile range – IQR) and categorical variables between clinical groups (Wilcoxon and Fisher’s exact tests, respectively). For microbial community profiling, we included samples that produced >300 high quality microbial reads for both 16S-Seq and Nanopore sequencing. We performed alpha diversity (Shannon index) calculations for each available sample, and then conducted between group comparisons of alpha diversity with non-parametric tests to draw inferences on systematic differences of alpha diversity between groups as a measure of relative community fitness.^1^ We conducted beta diversity analyses (Manhattan distances, analyzed via permutation analysis of variance and visualized via principal coordinates analyses) with the R *vegan* and *mia* packages.^46^ We examined for differentially abundant taxa between groups following centered log-ratio (CLR) transformations with the *limma* package to fit weighted linear regression models, perform tests based on an empirical Bayes moderated *t*-statistic and obtain False Discovery Ratio corrected p-values.

We then examined the discovered bacterial taxa at genus level and classified them by two different classification schemes with clinical relevance^12^:

A. By oxygen requirements for bacterial metabolism:
  1. Obligate aerobes (referred to throughout as aerobes): bacteria that require oxygen to grow and survive, as they use oxygen as final electron acceptor in their respiratory chain.
  2. Obligative anaerobes (referred to throughout as anaerobes): bacteria that are unable to grow in the presence of oxygen, as they are unable to use oxygen as a final electron acceptor and are killed in the presence of oxygen.
  3. Facultative anaerobes: bacteria that can grow in the presence or absence of oxygen. They can use both aerobic and anaerobic respiration, depending on the availability of oxygen in their environment, switching from aerobic to anaerobic metabolism.
  4. Microaerophiles: bacteria that require a low level of oxygen to grow and survive, as they can grow at oxygen concentrations lower than those required by obligate aerobes but higher than those tolerated by obligate anaerobes.
  5. Variable: genera that included both aerobes and anaerobes and could not be classified further with confidence.
  6. Unclassifiable: taxa that were not classified at the genus or family level with confidence to allow assessment of their metabolic needs.
B. By pathogenicity for LRT infections:
  1. Common respiratory pathogens: bacteria considered to be typical pathogens when isolated in LRT microbiologic cultures.
  2. Oral-origin commensal bacteria: bacterial taxa that have been characterized as typical members of the lung microbiome in health and originate from the oral cavity.
  3. Other: taxa with unclear clinical significance that do not fall into categories B1 or B2 above.

To agnostically examine our samples for distinct clusters of microbial composition (“metacommunities”), we applied unsupervised Dirichlet multinomial models (DMMs) with Laplace approximations^18^ to define the optimal number of clusters in our dataset, and then examined for associations with clinical parameters and outcomes. To synthesize bacterial and fungal data within each compartment, as well as bacterial profiles across different compartments, we used the weighted Similarity Network Fusion function.^19^ We classified subjects into a hyper-vs. hypo-inflammatory subphenotype based on predictions from a parsimonious logistic regression model utilizing plasma levels of sTNFR1, Ang-2 and procalcitonin (research biomarkers measured with Luminex panel), as well as serum bicarbonate levels measured during clinical care.

We followed patients prospectively and constructed Kaplan-Meier curves and Cox-proportional hazard models for 60-day survival, adjusted for the predictors of age and sex, as well as plausible confounders of microbiome associations diagnosis based on our findings (history of COPD, history of Immunosuppression), severity of illness as per the SOFA score, and host-response subphenotypes. To examine for the impact of mechanical ventilation, steroids and antibiotics pressure on longitudinal microbiota profiles, we constructed mixed regression models with random patient intercepts and adjusted for the number of days post-intubation that each sample was taken (as a proxy for the exposure to the hyperoxic environment of the ventilator) and the antibiotic exposure and steroids metrics by the day of sampling. We performed all statistical analyses in R v.4.2.0.^47^

Following derivation of the DMM clusters in each compartment of the UPMC-ARF cohort and demonstration of significant associations with patient outcomes, we proceeded to develop multinominal logistic regression models for prediction of classification of bacterial 16S profiles from new samples into predicted cluster assignments. We considered these new classification models as a Dysbiosis Index for each compartment. To develop these models in each compartment (oral, lung and gut), we used probabilistic graphical modeling (PGM)^48^ by considering the 50 most abundant taxa in each compartment along with the Shannon Index. We divided the samples of each compartment into two random subsets: 80% of data points for training and 20% for testing. The training set was used to generate a PGM using the FCI-MAX algorithm with Alpha of 0.1 to examine which variables (50 taxa abundance and Shannon Index) were associated with the cluster assignments in each compartment. The variables that appeared in the Markov blanket of the DMM cluster assignment variable were used to create a multinomial logistic regression (MLR) model to predict the cluster assignment of future samples. The MLR model equations were written as follows for the different cluster assignments (Low, Intermediate and High Diversity):

## Model equations

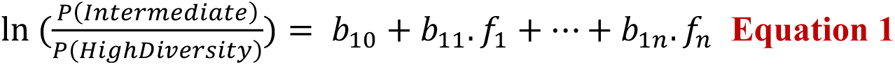

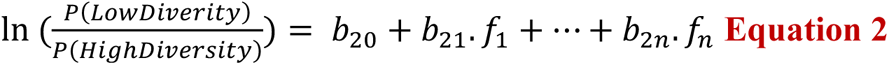

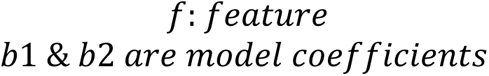

By rewriting the equations, we get the following:

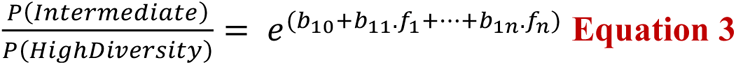

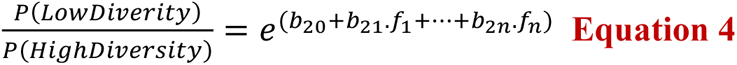

We rewrote the names of the model parameters as:

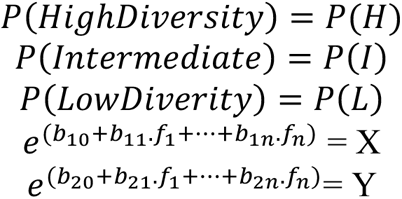

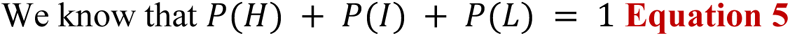

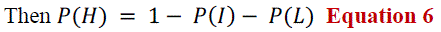

From **Equation 3 and 4**

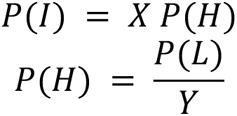

Substituting in **Equation 6**

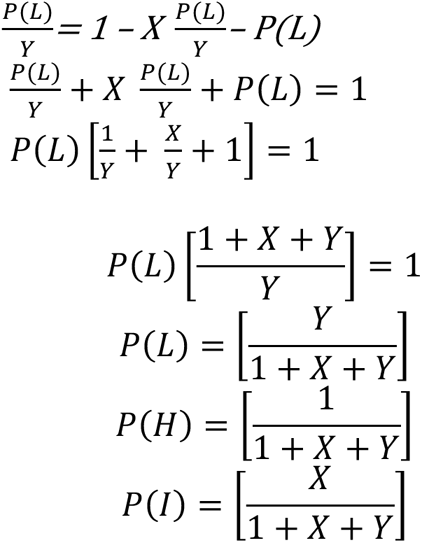

The predicted cluster is the one with the highest probability. For example, if *max*(*P*(*H*), *P*(*I*), *P*(*L*)) = *P*(*I*), then the predicted cluster is *P*(*I*)

The Intercepts and co-efficients for the MLR models for each compartment are provided below.

| Oral |  |  |
| --- | --- | --- |
| $f$ | $b_1$ | $b_2$ |
| Intercept | 8.124887 | 13.323657 |
| ShannonIndex | -1.57054 | -2.79173 |
| Actinomyces | 0.4883585 | 0.1975639 |
| Capnocytophaga | 0.1550266 | -0.2497556 |
| Fusobacterium | 0.04705918 | -0.49428753 |
| Granulicatella | 0.03521261 | -0.22396119 |
| Leptotrichia | -0.5982385 | -0.6815710 |
| Parvimonas | -0.8593016 | -0.6830944 |
| Porphyromonas | -0.7968399 | -0.4426564 |
| Prevotellaceae_uncl | -0.3983984 | -1.9055700 |
| Stomatobaculum | -0.5966283 | -1.3445377 |

| Lung |  |  |
| --- | --- | --- |
| $f$ | $b_1$ | $b_2$ |
| Intercept | 6.091456 | 13.161966 |
| ShannonIndex | -2.235179 | -3.955922 |
| Streptococcus | 0.08810083 | -0.30461314 |
| Veillonella | 0.27052525 | 0.04224102 |
| Peptostreptococcus | -0.02538055 | 0.50470722 |
| Porphyromonas | -0.520896 | -1.387346 |
| Selenomonas | -0.5639481 | -1.2279826 |
| Alloprevotella | -0.4193465 | -1.0781599 |
| Leptotrichia | -0.09918638 | -1.16197619 |
| Neisseriaceae_uncl | -0.430702 | -1.654160 |

| Gut |  |  |
| --- | --- | --- |
| $f$ | $b_1$ | $b_2$ |
| Intercept | -0.7901505 | 4.5913451 |
| ShannonIndex | 2.104042 | -1.017060 |
| Atopobium | 1.0404839 | 0.1382178 |
| Streptococcus | -0.1680732 | 0.2080845 |
| Enterococcus | -0.04209851 | 0.38397607 |
| Faecalibacterium | -0.8249428 | -0.9945793 |
| Fenollaria | 0.4178598 | -0.5862931 |
| Alistipes | -1.3291409 | -0.6846156 |
| Lachnospiraceae_uncl | -1.1289312 | -0.7073528 |

The Intercepts and co-efficients for the MLR models for each compartment are provided below.

## Data Availability

Data and code availability:
Sequencing data collected for the study have been deposited to the Sequencing Resource Archive, through the following Accession numbers:
-PRJNA595346 for 16S data of UPMC-ARF and UPMC-COVID cohorts,
-PRJNA726955 for ITS data of UPMC-ARF cohort,
-PRJNA554461 for Nanopore data of UPMC-ARF cohort,
-PRJNA940725 for 16S data of the Healthy Controls,
-PRJNA976404 for Metagenomic data of the MGH-COVID cohort.
Primary code and de-identified data for replication of analyses will be available on the github repository (https://github.com/MicrobiomeALIR/MultiCompartmentMicrobiome) upon acceptable of the manuscript for publication. Any additional information required to reanalyze the data reported in this paper is available from the lead contact upon request.

## Acknowledgements

The authors wish to thank the patients and patient families that have enrolled in the University of Pittsburgh Acute Lung Injury Registry. We also thank the physicians, nurses, respiratory therapists and other staff at the University of Pittsburgh Medical Center Presbyterian, Shadyside and East Hospitals intensive care units for assistance with coordination of patient enrollment and collection of patient samples. We would like to thank the laboratory personnel at the Center for Medicine and the Microbiome at the University of Pittsburgh for assistance with processing clinical samples. We acknowledge the contributions of Nameer Al-Yousif, MD, Michael Lu, MD, Grace Lisius MD, and Caitlin Shaefer, MPH who participated in clinical data extractions for specific components of the databases of the UPMC-ARF and UPMC-COVID cohorts. We also thank the Massachusetts General Hospital Translational and Clinical Research Center (TCRC) for their support of the project and the assembly of the MGH-COVID cohort.

## Ethics approval and consent to participate

The University of Pittsburgh Institutional Review Board (IRB) approved the protocol for the UPMC-ARF and UPMC-COVID cohorts (STUDY19050099). We obtained written or electronic informed consent by all participants or their surrogates in accordance with the Declaration of Helsinki. For the MGH-COVID cohort, the Study protocol #2020P000804 was approved by the Mass General Brigham IRB. For the healthy controls, the University of Pittsburgh IRB approved the study protocols (STUDY19060243 for respiratory biospecimens and STUDY20060312 for stool biospecimens). All participants or their healthcare proxy provided written informed consent to participate.

## Consent for publication

We obtained necessary patient/participant consent and the appropriate institutional forms have been archived. Any patient/participant/sample identifiers included were not known to anyone outside the research group so cannot be used to identify individuals.

## Data and code availability

Sequencing data collected for the study have been deposited to the Sequencing Resource Archive, through the following Accession numbers:

– PRJNA595346 for 16S data of UPMC-ARF and UPMC-COVID cohorts (477 records released and remainder to be released upon publication with Temporary Submission ID SUB13319619),
– PRJNA726955 for ITS data of UPMC-ARF cohort,
– PRJNA554461 for Nanopore data of UPMC-ARF cohort,
– PRJNA940725 for 16S data of the Healthy Controls,
– PRJNA976404 for Metagenomic data of the MGH-COVID cohort.

Primary code and de-identified data for replication of analyses will be available on the github repository (https://github.com/MicrobiomeALIR/MultiCompartmentMicrobiome) upon acceptable of the manuscript for publication. Any additional information required to reanalyze the data reported in this paper is available from the lead contact upon request.

## REFERENCES

1. Kitsios, G. D. et al. Dysbiosis in the intensive care unit: Microbiome science coming to the bedside. J. Crit. Care 38, 84–91 (2017).

2. Dickson, R. P. The microbiome and critical illness. Lancet Respir. Med. 4, 59–72 (2016).

3. Kitsios, G. D. et al. Respiratory Tract Dysbiosis Is Associated with Worse Outcomes in Mechanically Ventilated Patients. Am. J. Respir. Crit. Care Med. 202, 1666–1677 (2020).

4. Dickson, R. P. et al. Lung microbiota predict clinical outcomes in critically ill patients. Am. J. Respir. Crit. Care Med. 201, 555–563 (2020).

5. Sulaiman, I. et al. Microbial signatures in the lower airways of mechanically ventilated COVID-19 patients associated with poor clinical outcome. Nat. Microbiol. 6, 1245–1258 (2021).

6. Sarma, A., Calfee, C. S. & Ware, L. B. Biomarkers and precision medicine: state of the art. Crit. Care Clin. 36, 155–165 (2020).

7. Kitsios, G. D. et al. Host-Response Subphenotypes Offer Prognostic Enrichment in Patients With or at Risk for Acute Respiratory Distress Syndrome. Crit. Care Med. 47, 1724–1734 (2019).

8. Alipanah, N. & Calfee, C. S. Phenotyping in acute respiratory distress syndrome: state of the art and clinical implications. Curr. Opin. Crit. Care 28, 1–8 (2022).

9. Heijnen, N. F. L. et al. Biological Subphenotypes of Acute Respiratory Distress Syndrome Show Prognostic Enrichment in Mechanically Ventilated Patients without Acute Respiratory Distress Syndrome. Am. J. Respir. Crit. Care Med. 203, 1503–1511 (2021).

10. Kitsios, G. D. et al. Distinct profiles of host responses between plasma and lower respiratory tract during acute respiratory failure. ERJ Open Research 9, (2023).

11. Fair, K., et al. Rectal Swabs from Critically Ill Patients Provide Discordant Representations of the Gut Microbiome Compared to Stool Samples. mSphere 4, (2019).

12. Kitsios, G. D. et al. The upper and lower respiratory tract microbiome in severe aspiration pneumonia. iScience 26, 106832 (2023).

13. Britton, N. et al. Respiratory Fungal Communities are Associated with Systemic Inflammation and Predict Survival in Patients with Acute Respiratory Failure. medRxiv (2023) doi:10.1101/2023.05.11.23289861.

14. ARDS Definition Task Force et al. Acute respiratory distress syndrome: the Berlin Definition. JAMA 307, 2526–2533 (2012).

15. Nguyen, L. H. et al. Metagenomic assessment of gut microbial communities and risk of severe COVID-19. Genome Med. 15, 49 (2023).

16. Zhao, J., Murray, S. & Lipuma, J. J. Modeling the impact of antibiotic exposure on human microbiota. Sci. Rep. 4, 4345 (2014).

17. Wang, A. A. et al. The Narrow-Spectrum Antibiotic Treatment Score: A Novel Quantitative Tool for Assessing Broad– and Narrow-Spectrum Antibiotic Use in Severe Community-Acquired Pneumonia. in *B28*. HOST AND MICROBIAL CLINICAL STUDIES IN LUNG INFECTIONS AND LUNG DISEASES A2929–A2929 (American Thoracic Society, 2020). doi:10.1164/ajrccm-conference.2020.201.1_MeetingAbstracts.A2929.

18. Holmes, I., Harris, K. & Quince, C. Dirichlet multinomial mixtures: generative models for microbial metagenomics. PLoS ONE 7, e30126 (2012).

19. Narayana, J. K., Mac Aogáin, M., Ali, N. A. B. M., Tsaneva-Atanasova, K. & Chotirmall, S. H. Similarity network fusion for the integration of multi-omics and microbiomes in respiratory disease. Eur. Respir. J. 58, (2021).

20. Drohan, C. M. et al. Biomarker-Based Classification of Patients With Acute Respiratory Failure Into Inflammatory Subphenotypes: A Single-Center Exploratory Study. Crit. Care Explor. 3, e0518 (2021).

21. Kitsios, G. D., Franz, C. & McVerry, V. The Microbiome in Acute Lung Injury and ARDS. in The Microbiome in Respiratory Disease (eds. Huang, Y. J. & Garantziotis, S.) (Humana, Cham, 2022).

22. Lloréns-Rico, V. et al. Clinical practices underlie COVID-19 patient respiratory microbiome composition and its interactions with the host. Nat. Commun. 12, 6243 (2021).

23. Bernard-Raichon, L. et al. Gut microbiome dysbiosis in antibiotic-treated COVID-19 patients is associated with microbial translocation and bacteremia. Nat. Commun. 13, 5926 (2022).

24. Chanderraj, R. et al. In critically ill patients, anti-anaerobic antibiotics increase risk of adverse clinical outcomes. Eur. Respir. J. (2022) doi:10.1183/13993003.00910-2022.

25. Pickens, C. O. et al. Bacterial Superinfection Pneumonia in Patients Mechanically Ventilated for COVID-19 Pneumonia. Am. J. Respir. Crit. Care Med. 204, 921–932 (2021).

26. Kullberg, R. F. J., Schinkel, M. & Wiersinga, W. J. Empiric anti-anaerobic antibiotics are associated with adverse clinical outcomes in emergency department patients. Eur. Respir. J. 61, (2023).

27. Kitsios, G. D. & McVerry, B. J. Host-Microbiome Interactions in the Subglottic Space. Bacteria Ante Portas! Am. J. Respir. Crit. Care Med. 198, 294–297 (2018).

28. Dickson, R. P. et al. Bacterial topography of the healthy human lower respiratory tract. MBio 8, (2017).

29. Zhao, T. et al. Oral hygiene care for critically ill patients to prevent ventilator-associated pneumonia. Cochrane Database Syst. Rev. 12, CD008367 (2020).

30. Hammond, N. E. et al. Association Between Selective Decontamination of the Digestive Tract and In-Hospital Mortality in Intensive Care Unit Patients Receiving Mechanical Ventilation: A Systematic Review and Meta-analysis. JAMA 328, 1922–1934 (2022).

31. Klompas, M. Oropharyngeal Decontamination with Antiseptics to Prevent Ventilator-Associated Pneumonia: Rethinking the Benefits of Chlorhexidine. Semin. Respir. Crit. Care Med. 38, 381–390 (2017).

32. Buelow, E. et al. Comparative gut microbiota and resistome profiling of intensive care patients receiving selective digestive tract decontamination and healthy subjects. Microbiome 5, 88 (2017).

33. Horn, K. J., Schopper, M. A., Drigot, Z. G. & Clark, S. E. Airway Prevotella promote TLR2-dependent neutrophil activation and rapid clearance of Streptococcus pneumoniae from the lung. Nat. Commun. 13, 3321 (2022).

34. Segal, L. N. et al. Enrichment of lung microbiome with supraglottic taxa is associated with increased pulmonary inflammation. Microbiome 1, 19 (2013).

35. Wu, B. G. et al. Episodic Aspiration with Oral Commensals Induces a MyD88-dependent, Pulmonary T-Helper Cell Type 17 Response that Mitigates Susceptibility to Streptococcus pneumoniae. Am. J. Respir. Crit. Care Med. 203, 1099–1111 (2021).

36. Dickson, R. P. et al. Enrichment of the lung microbiome with gut bacteria in sepsis and the acute respiratory distress syndrome. Nat. Microbiol. 1, 16113 (2016).

37. Nath, S., Kitsios, G. D. & Bos, L. D. J. Gut-lung crosstalk during critical illness. Curr. Opin. Crit. Care 29, 130–137 (2023).

38. Bain, W. et al. Research Bronchoscopy Standards and the Need for Non-Invasive Sampling of the Failing Lungs. Ann Am Thorac Soc (2023).

39. Shankar-Hari, M., Fan, E. & Ferguson, N. D. Acute respiratory distress syndrome (ARDS) phenotyping. Intensive Care Med. 45, 516–519 (2019).

40. Kalil, A. C. et al. Management of Adults With Hospital-acquired and Ventilator-associated Pneumonia: 2016 Clinical Practice Guidelines by the Infectious Diseases Society of America and the American Thoracic Society. Clin. Infect. Dis. 63, e61–e111 (2016).

41. Morris, A. et al. Comparison of the respiratory microbiome in healthy nonsmokers and smokers. Am. J. Respir. Crit. Care Med. 187, 1067–1075 (2013).

42. Kitsios, G. D. et al. Respiratory microbiome profiling for etiologic diagnosis of pneumonia in mechanically ventilated patients. Front. Microbiol. 9, 1413 (2018).

43. Charalampous, T. et al. Nanopore metagenomics enables rapid clinical diagnosis of bacterial lower respiratory infection. Nat. Biotechnol. 37, 783–792 (2019).

44. Yang, L. et al. Metagenomic identification of severe pneumonia pathogens in mechanically-ventilated patients: a feasibility and clinical validity study. Respir. Res. 20, 265 (2019).

45. Juul, S., et al. What’s in my pot? Real-time species identification on the MinION. BioRxiv (2015) doi:10.1101/030742.

46. Huber, W. et al. Orchestrating high-throughput genomic analysis with Bioconductor. Nat. Methods 12, 115–121 (2015).

47. R Foundation for Statistical Computing, R. C. T. R: A Language and Environment for Statistical Computing. (CRAN, 2016).

48. Raghu, V. K. et al. Comparison of strategies for scalable causal discovery of latent variable models from mixed data. Int. J. Data Sci. Anal. 6, 33–45 (2018).

