## Supplement for "Prognostic Insights from Longitudinal Multicompartment Study of Host-Microbiota Interactions in Critically Ill Patients"

**Extended Data Figures and Tables:**

**Table S1: Clinical characteristics of subjects enrolled in the COVID-19 cohorts.**

| | UPMC-COVID | MGH-COVID |
|---|---|---|
| N | 49 | 97 |
| Age, years (median [IQR]) | 63.0 [57.1, 71.7] | 63.0 [51.0, 71.0] |
| Men, n (%) | 30 (61.2) | 54 (55.7) |
| Whites, n (%) | 37 (75.5) | 70 (72.2) |
| BMI (median [IQR]) | 33.9 [29.0, 41.8] | 30.1 [26.6, 36.0] |
| COPD, n (%) | 10 (20.4) | 38 (39.2) |
| Diabetes, n (%) | 21 (42.9) | 30 (30.9) |
| WBC (median [IQR]) | 11.2 [8.6, 14.1] | NA |
| Creatinine (median [IQR]) | 1.0 [0.7, 1.6] | NA |
| Plateau Pressure (median [IQR]) | 26.0 [20.0, 29.0] | NA |
| PaO2:FiO2 ratio (median [IQR]) | 94.0 [69.8, 168.2] | NA |
| Hypoinflammatory subphenotype, n (%) | 42 (87.5) | NA |
| VFD (median [IQR]) | 0.0 [0.0, 13.0] | NA |
| Severe Disease, n (%) | 49 (100) | 41 (42.3) |
| Mortality 60-day, n (%) | 23 (46.9) | 10 (10.3) |

**Figure S1: Quality control steps for clinical versus experimental control samples.** A-B: Clinical samples from critically ill patients and healthy controls he had much higher sequencing yield (high quality reads from Illumina MiSeq 16S rRNA gene sequencing) and markedly different bacterial composition (as shown in Principal Coordinates Analysis) compared to negative controls or PCR positive samples. C-D: Quality control examination for gut samples. Unsoiled rectal swabs (i.e. not visibly coated by stool) had markedly lower bacterial burden (examined by qPCR of 16S rRNA gene) and differential composition compared to soiled rectal swabs or stool samples, and therefore, unsold rectal swabs were excluded from further analysis as they may not provide sufficient representation of gut microbiota.

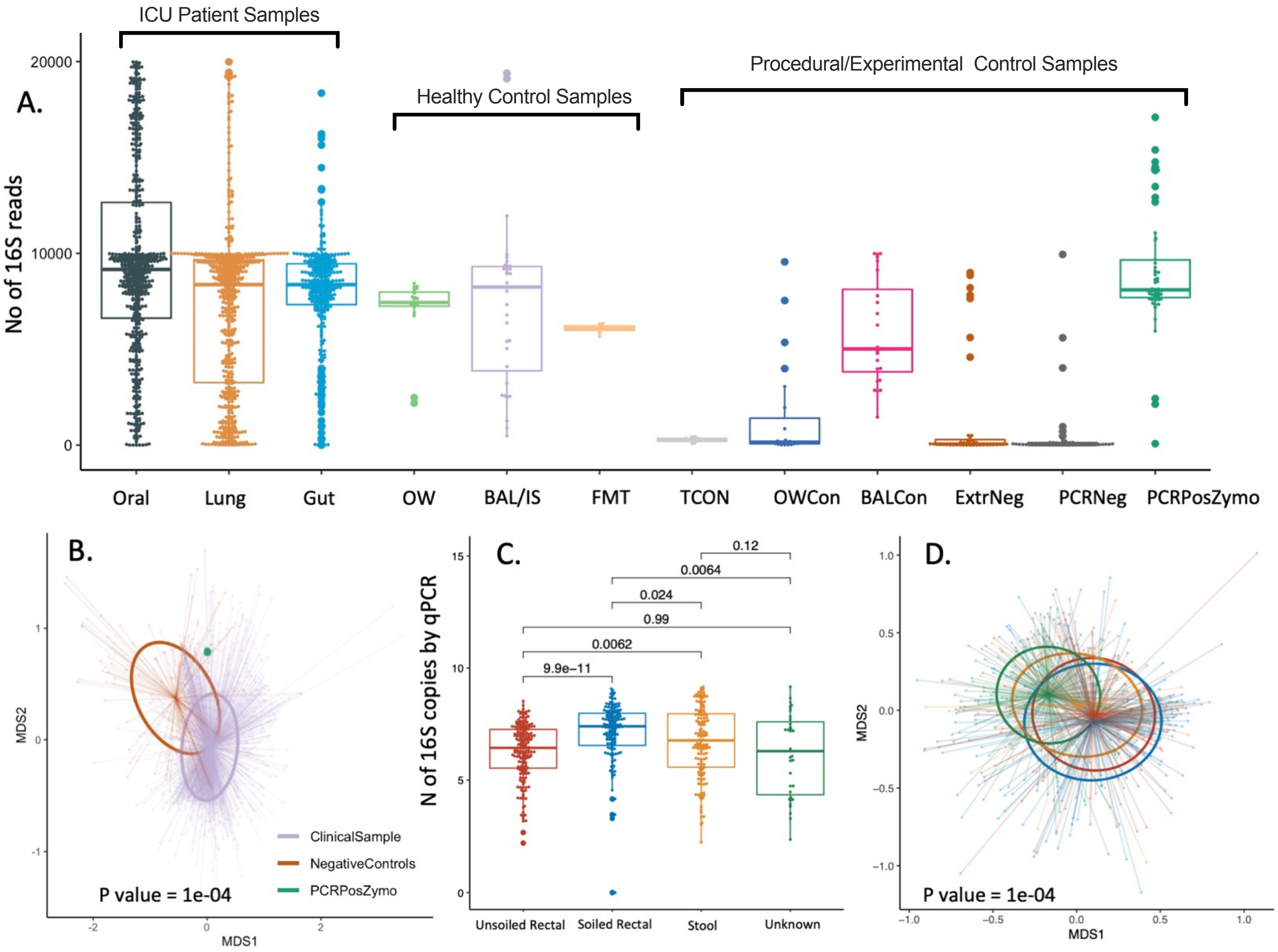

**Figure S2. Enrichment of respiratory track samples by gut origin bacteria.** A. Relative abundance of all available oral and lung samples for gut origin taxa (*Enterococcus, Enterobacteriaceae, Escherichia_Shigella, Klebsiella, Bacteroides, Lachonoclostridium, Lachnospiraceae_ge, Lachnospiraceae_uncl, Anaerococcus*) vs. other taxa. 4.8% and 8.1% of all oral and lung samples, respectively, had >30% relative abundance for gut-origin taxa (Fisher's p=0.03), classified as samples with Gut Enrichment. B. Proportion of oral lung samples with gut origin taxa enrichment across the three time intervals of sampling. There was increase in the proportion of lung samples with gut origin taxa enrichment from baseline to middle interval (Fisher test p=0.02). C-D. Relative abundance of individual gut origin taxa for the oral and lung samples with gut enrichment. Gut enrichment was mostly accounted for by organisms with pathogenic potential (*Enterococcus, Enterobacteriaceae, Escherichia_Shigella* and *Klebsiella*, shown in blue colors) than gut origin commensals (*Bacteroides, Lachonoclostridium, Lachnospiraceae_ge, Lachnospiraceae_uncl, Anaerococcus*). E-F. Patients with gut origin taxa enrichment in lung samples at baseline had significantly worse 60-day survival (log rank p<0.0001) compared to patients without enrichment for gut origin taxa, whereas gut enrichment in oral samples did not impact survival.

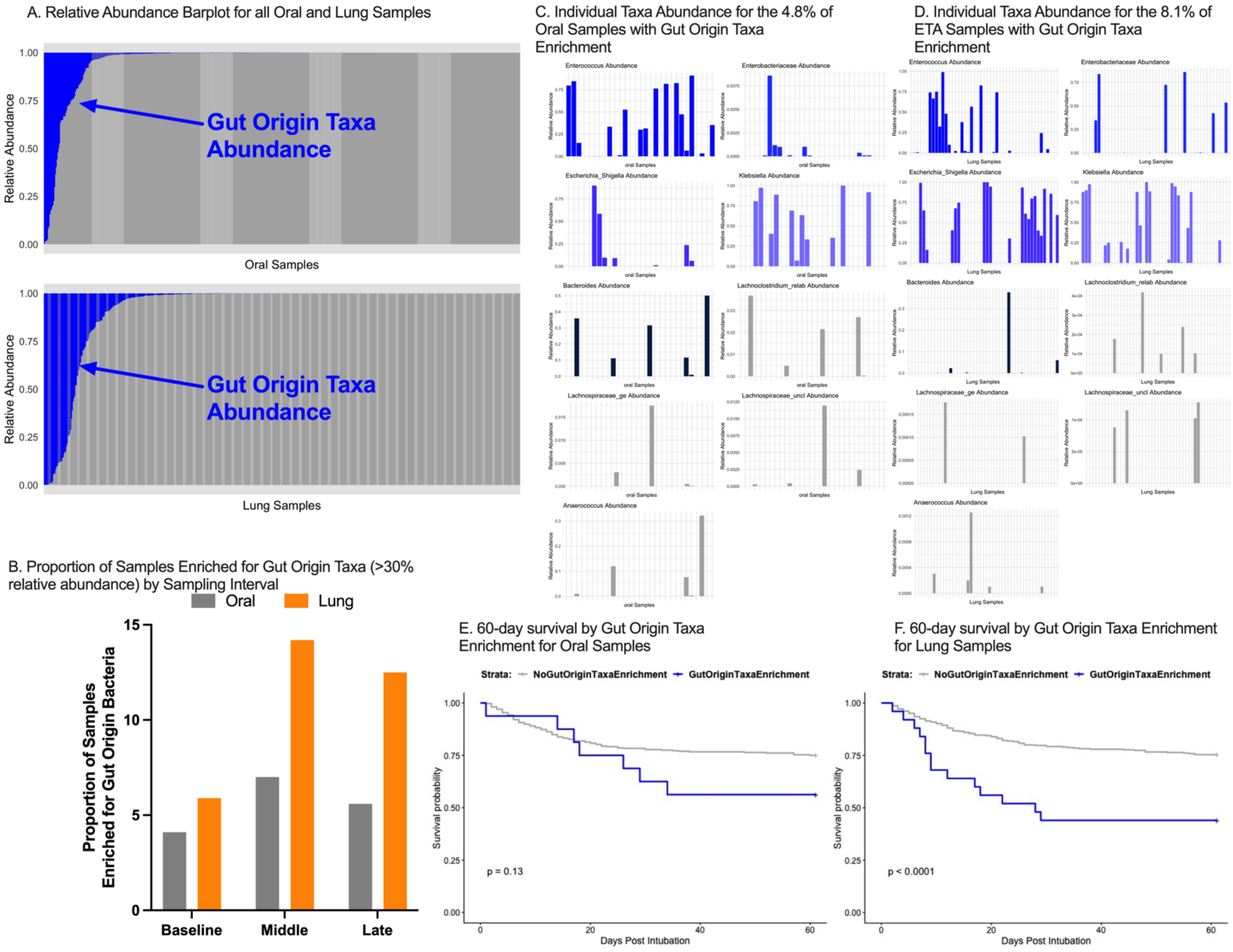

A. Relative Abundance Barplot for all Oral and Lung Samples
Relative Abundance
1.00
0.75
0.50
0.25
0.00
Gut Origin Taxa Abundance
Oral Samples
Gut Origin Taxa Abundance
Lung Samples
C. Individual Taxa Abundance for the 4.8% of Oral Samples with Gut Origin Taxa Enrichment
Enterococcus Abundance
Enterobacteriaceae Abundance
Escherichia_Shigella Abundance
Klebsiella Abundance
Bacteroides Abundance
Lachnoclostridium_relab Abundance
Lachnospiraceae_ge Abundance
Lachnospiraceae_uncl Abundance
Anaerococcus Abundance
oral Samples
D. Individual Taxa Abundance for the 8.1% of ETA Samples with Gut Origin Taxa Enrichment
Enterococcus Abundance
Enterobacteriaceae Abundance
Escherichia_Shigella Abundance
Klebsiella Abundance
Bacteroides Abundance
Lachnoclostridium_relab Abundance
Lachnospiraceae_ge Abundance
Lachnospiraceae_uncl Abundance
Anaerococcus Abundance
Lung Samples
B. Proportion of Samples Enriched for Gut Origin Taxa (>30% relative abundance) by Sampling Interval
Oral
Lung
Proportion of Samples Enriched for Gut Origin Bacteria
15
10
5
0
Baseline
Middle
Late
E. 60-day survival by Gut Origin Taxa Enrichment for Oral Samples
Strata: NoGutOriginTaxaEnrichment GutOriginTaxaEnrichment
Survival probability
1.00
0.75
0.50
0.25
0.00
p = 0.13
0
20
40
60
Days Post Intubation
F. 60-day survival by Gut Origin Taxa Enrichment for Lung Samples
Strata: NoGutOriginTaxaEnrichment GutOriginTaxaEnrichment
Survival probability
1.00
0.75
0.50
0.25
0.00
p < 0.0001
0
20
40
60
Days Post Intubation

**Figure S3: Fungal ITS sequencing and Nanopore metagenomic results.** ITS sequencing was performed in samples from all three compartments [oral (n=226), lung (n=287), gut (n=31)], whereas Nanopore metagenomics was performed in 130 lung samples. *C. albicans* was the most abundant fungus and dominated (>50% relative abundance) all three compartments at all time points (A-B). Oral and lung communities in follow-up samples had lower fungal Shannon index compared to baseline samples (B). History of immunosuppression and diagnosis of pneumonia were positively correlated with *C. albicans* abundance in oral and lung samples (C). Comparison of 16S and Nanopore derived lung communities (D-E) showed that *Streptococcus* was the most abundant taxon by both methodologies, whereas Nanopore metagenomics confirmed that *C.albicans* was the second most abundant taxon in lung communities.

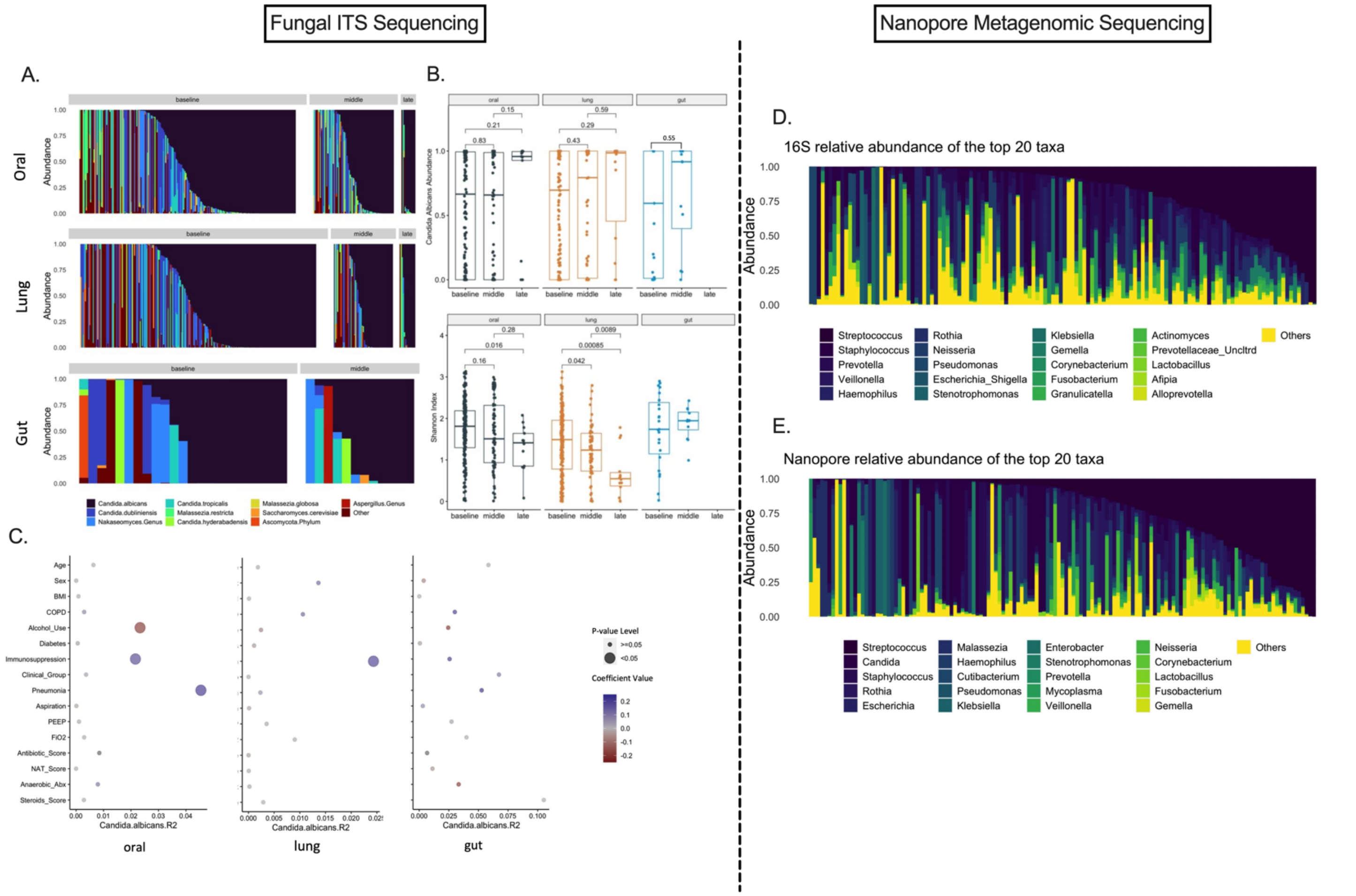

**Figure S4: Clinical variables associated with alpha diversity (Shannon Index), obligate anaerobe and respiratory pathogen abundance in baseline samples from the three body compartments.** Clinical variables are shown on the y-axis, and R2 of linear regression models for Shannon index (A), Anaerobe abundance (B) and Pathogen abundance (C) are shown on the x-axis. Statistically significant associations ($p<0.05$) are shown with large bubbles and direction of association is color coded.

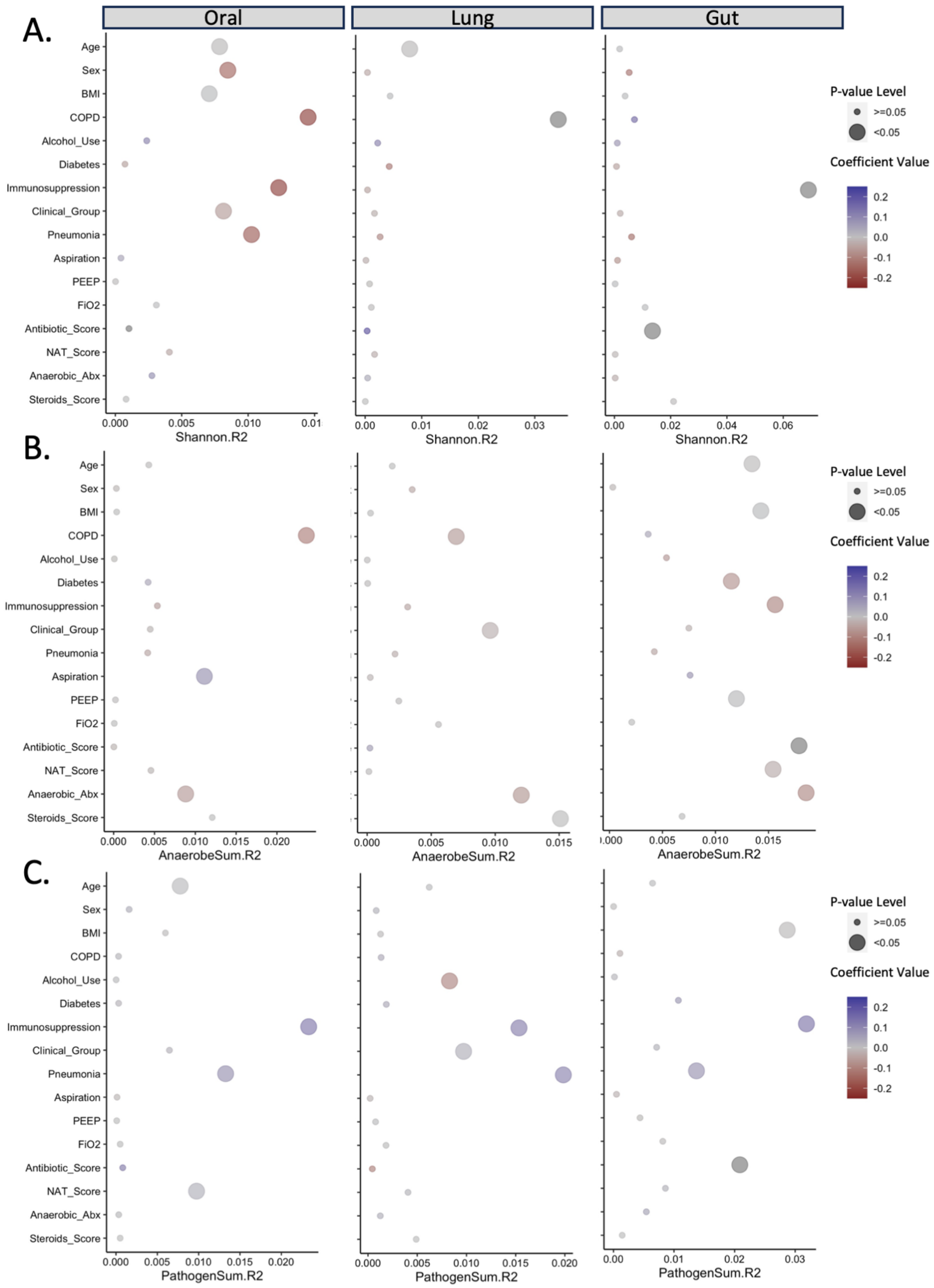

A.
Oral
Lung
Gut
Age
Sex
BMI
COPD
Alcohol_Use
Diabetes
Immunosuppression
Clinical_Group
Pneumonia
Aspiration
PEEP
FiO2
Antibiotic_Score
NAT_Score
Anaerobic_Abx
Steroids_Score
Shannon.R2
B.
AnaerobeSum.R2
C.
PathogenSum.R2
P-value Level
>=0.05
<0.05
Coefficient Value
0.2
0.1
0.0
-0.1
-0.2

**Table S2: Mixed linear regression models for the examination of the effects of antibiotics and steroids on features of dysbiosis in samples from all three compartments. We examined the antibiotic exposure coded in three different ways:** i) anaerobic coverage, ii) a numerical scale that included duration, timing and type, and iii) the Narrow Antibiotic Treatment (NAT) score. Each effect was adjusted for the study day from enrollment. The p-values of the mixed effects models with random patient intercepts are shown for each endpoint (columns) and significant values are highlighted in bold.

| **Variables** | **Shannon index** | **Obligate anaerobe abundance** | **Respiratory Pathogen abundance** |
|---|---|---|---|
| **Oral** | | | |
| Anaerobic_spectrum | 0.2119 | **0.0315** | 0.6208 |
| Antibiotic_score | 0.9415 | 0.6623 | 0.0717 |
| NAT_score | 0.4152 | 0.1868 | **0.0386** |
| Steroids_score | 0.7290 | 0.0515 | 0.5649 |
| **Lung** | | | |
| Anaerobic_spectrum | 0.7660 | **0.0287** | 0.4178 |
| Antibiotic_score | 0.8019 | 0.1841 | 0.1788 |
| NAT_score | 0.5088 | 0.8518 | 0.2759 |
| Steroids_score | 0.8010 | **0.027** | 0.3364 |
| **Gut** | | | |
| Anaerobic_spectrum | 0.5029 | **0.0119** | 0.0977 |
| Antibiotic_score | 0.207 | 0.1881 | **0.0236** |
| NAT_score | 0.6993 | **0.0104** | **0.0396** |
| Steroids_score | 0.0911 | 0.3017 | 0.6127 |

**Figure S5: Longitudinal analysis of bacterial DMM cluster assignments showed overall stability from baseline to middle interval for Low-Diversity samples in each compartment.** Low-Diversity cluster is shown in brown, Intermediate-Diversity in light red, and High-Diversity in blue. Only subjects with available samples on both baseline and middle intervals are included in this analysis.

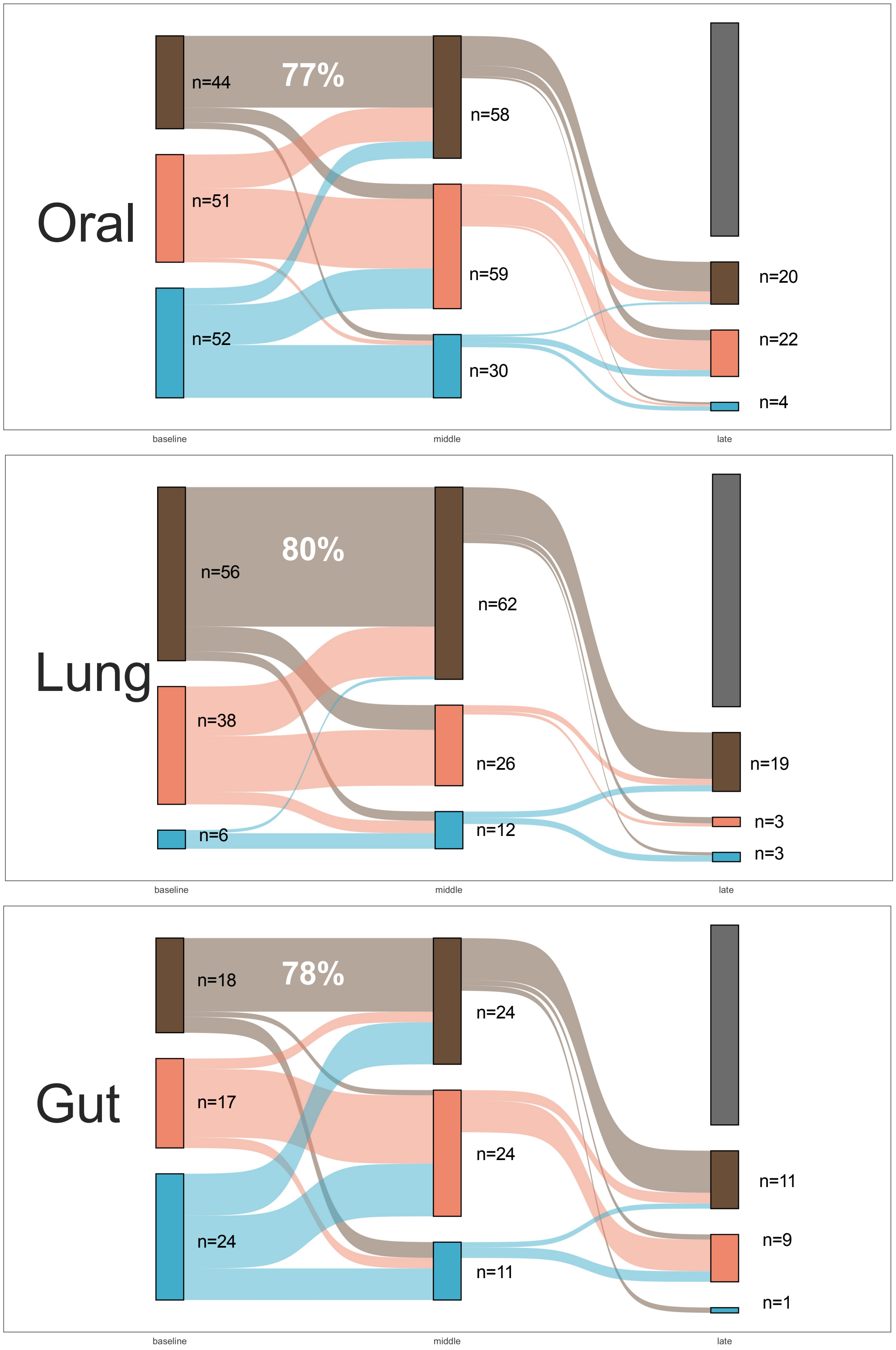

**Figure S6: Bacterial-Fungal SNF Clusters.** Comparisons of Shannon index, obligate anaerobe, pathogen and *C.albicans* abundance between clusters at each body compartment.

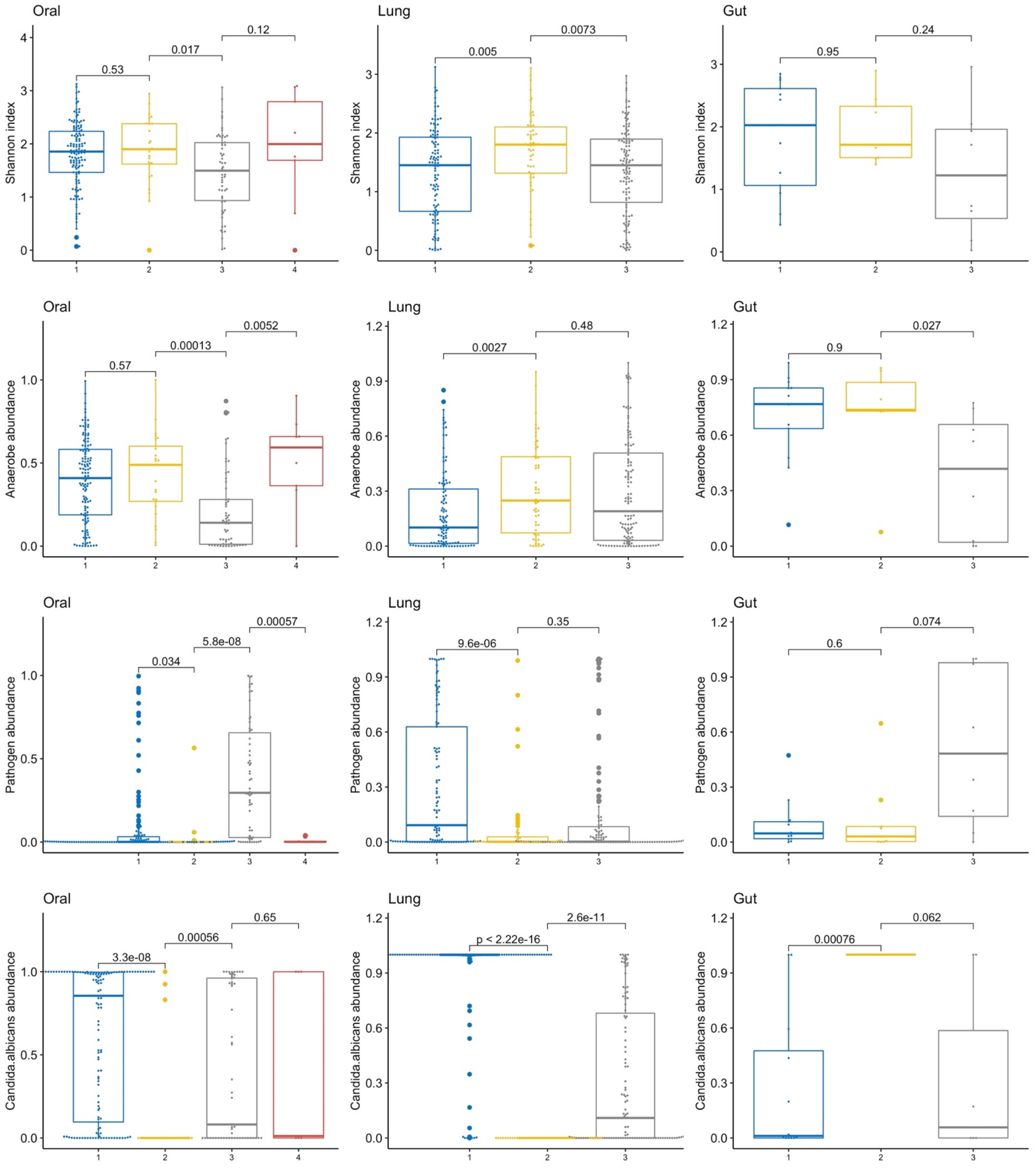

**Figure S7. Microbiota correlate with host response biomarkers at both the lung compartment and at a systemic level.** A-C: Heatmaps of correlations between the top 20 abundant taxa in the oral (A), lung (B) and gut (C) compartment with 10 host response biomarkers measured in plasma samples (top 10 rows) and endotracheal aspirate (ETA) supernatant samples (bottom 10 rows) in each heatmap. ETA biomarker values were adjusted for total protein concentration in each sample. Statistically significant correlations adjusted for multiple testing (Benjamini-Hochberg method) are shown with crosses ("+") and the direction of the correlation is color coded. D-E: Comparisons of ETA and plasma biomarkers between clusters: bacterial DMM clusters (D), Nanopore DMM clusters (E), and Bacterial-Fungal SNF clusters (F). The low diversity bacterial DMM cluster (D-brown) had significantly higher levels of plasma sTNFR1 and sRAGE and procalcitonin levels, whereas the low diversity Nanopore DMM cluster (E-orange) had higher levels of ETA IL-6, ETA sRAGE, plasma IL-6, plasma sTNFR1, plasma sRAGE, plasma Ang-2 and plasma Pentraxin-3. G. Chord plots for plasma-derived subphenotypes of host response (hyper- vs. hypo-inflammatory as predicted by a 4-variable regression model) and bacterial DMM clusters. No significant associations were found. H. Hyper-inflammatory patients had higher abundance of pathogens in lung samples compared to hypo-inflammatory patients but only among patients without diagnosis of pneumonia.

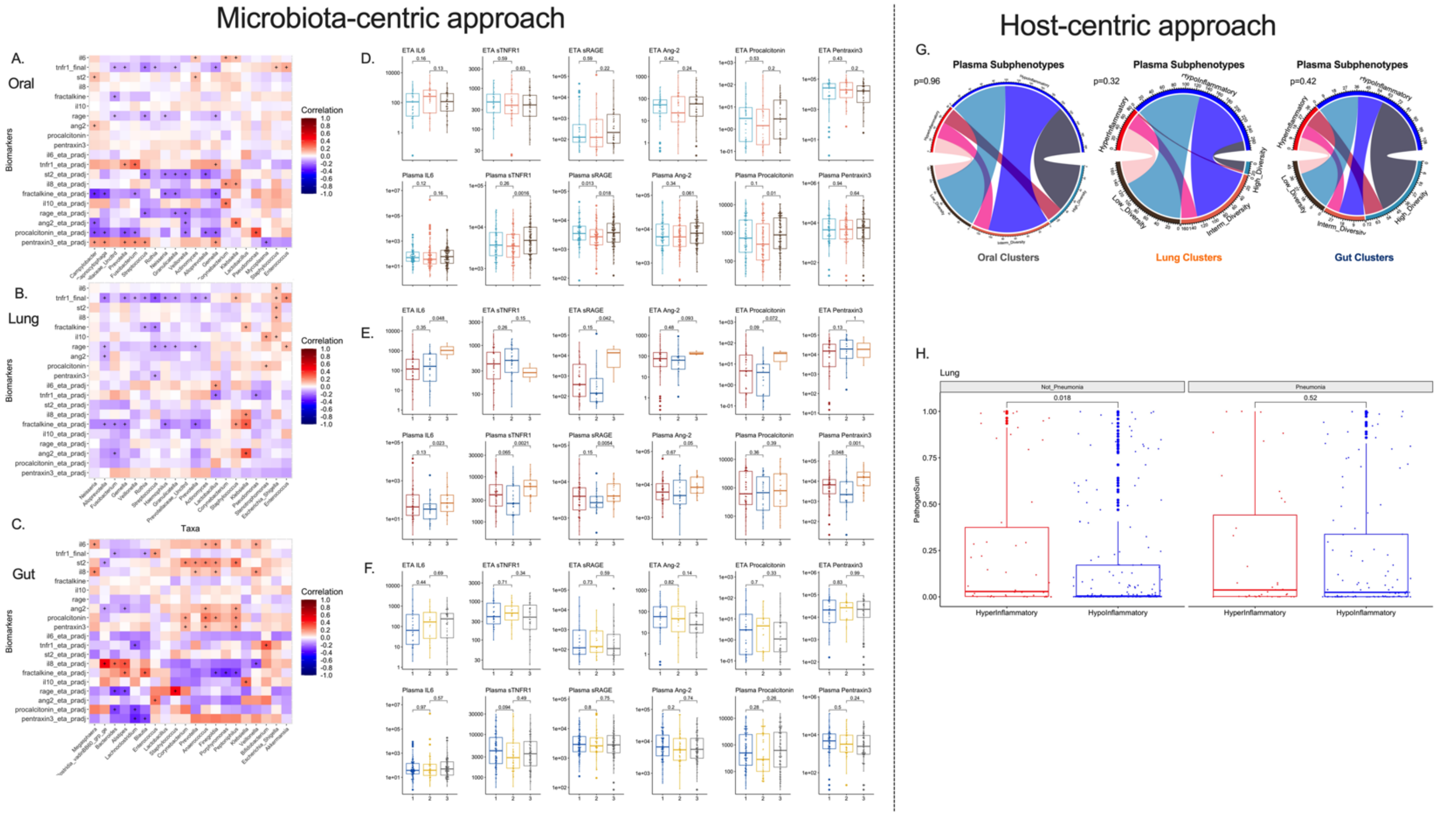

**Figure S8. Microbiota differences between 60-day survivors and non-survivors in the oral, lung and gut compartments.** Longitudinal differences in Shannon index (A), anaerobe abundance (B) and pathogen abundance (C) between survivors and non-survivors in each compartment.

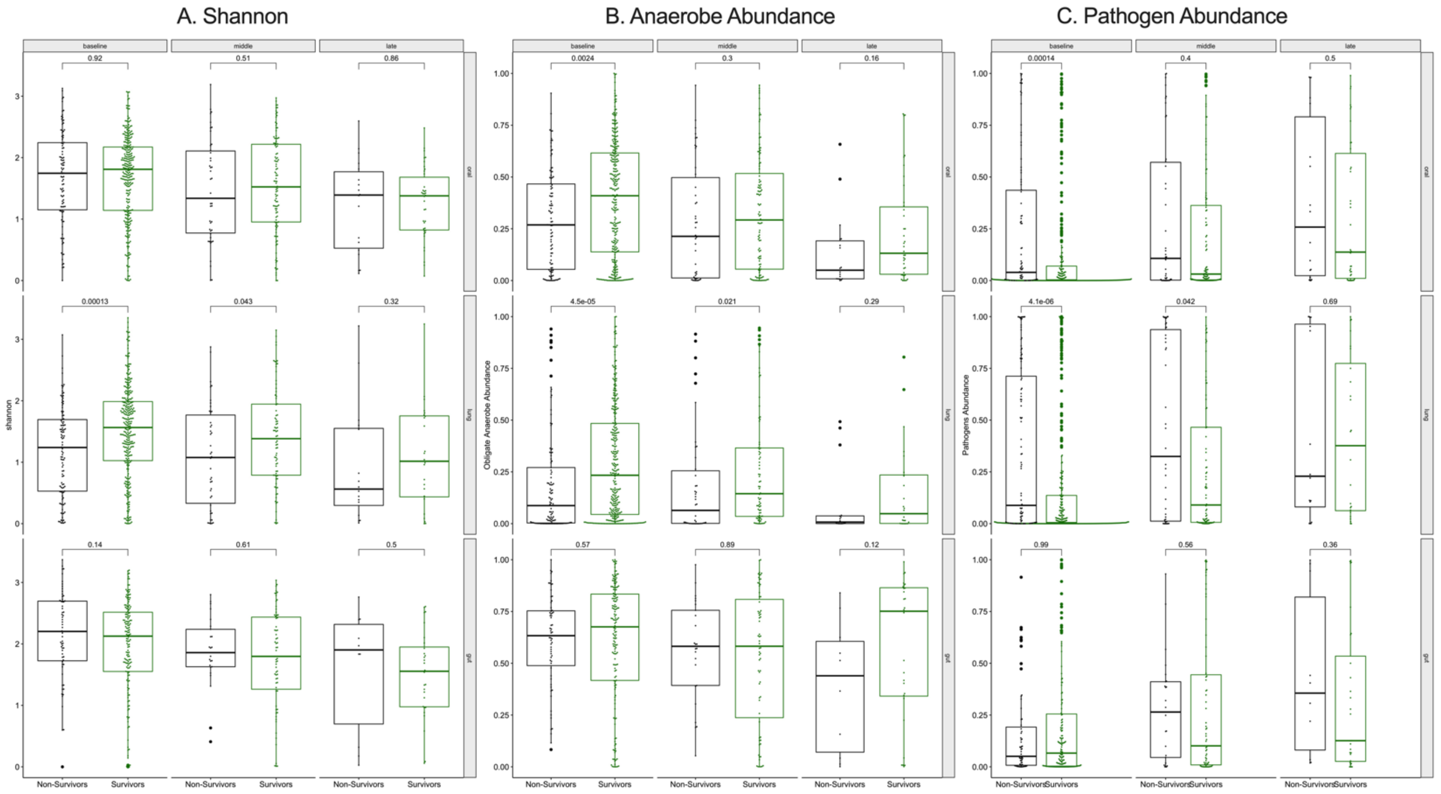
